# A rare gain of function HCN4 gene mutation is responsible for inappropriate sinus tachycardia in a Spanish family

**DOI:** 10.1101/2023.01.20.23284606

**Authors:** Anabel Cámara-Checa, Francesca Perin, Marcos Rubio-Alarcón, María Dago, Teresa Crespo-García, Josu Rapún, María Marín, Jorge Cebrián, Francisco Bermúdez-Jiménez, Lorenzo Monserrat, Juan Tamargo, Ricardo Caballero, Juan Jiménez-Jáimez, Eva Delpón

## Abstract

**Background:** In a family with inappropriate sinus tachycardia (IST) we identified a novel mutation (p.V240M) of the hyperpolarization-activated cyclic nucleotide-gated type 4 (HCN4) channel, which contributes to the pacemaker current (I_f_) in human sinoatrial node cells. Here we clinically study the family and functionally analyze the p.V240M variant.

**Methods:** Macroscopic (I_HCN4_) and single-channel currents were recorded using patch-clamp in cells expressing human native (WT) and/or p.V240M HCN4 channels.

**Results:** All p.V240M mutation carriers exhibited IST (mean heart rate 113[7] bpm, n=9), that in adults, was accompanied by cardiomyopathy. I_HCN4_ generated by p.V240M channels either alone or in combination with WT was significantly greater than that generated by WT channels. The variant, which lies in the N-terminal HCN domain, increased single-channel conductance and opening frequency and probability of HCN4 channels. Conversely, it did not modify channel sensitivity for cAMP and ivabradine or the level of expression at the membrane. Treatment with ivabradine based on functional data reversed the IST and the cardiomyopathy of the carriers.

**Conclusions:** The p.V240M gain-of-function variant increases I_f_ during diastole, which explains the IST of the carriers. The results demonstrate the importance of the unique HCN domain in HCN4 which stabilizes the channels in the closed state.

**Funding:** Ministerio de Ciencia e Innovación (PID2020-118694RB-I00); Comunidad Autónoma de Madrid (P2022/BMD-7229), European Structural and Investment Funds); and Instituto de Salud Carlos III (CIBERCV; CB16/11/00303).

## INTRODUCTION

Inappropriate sinus tachycardia (IST) is a clinical syndrome with an estimated prevalence of around 1% (more frequent in young women) and a significant impact on the quality of life of the affected individuals (Baruscotti et al., 2016; Olshansky and Sullivan, 2019). IST is defined by a sinus heart rate (HR) inexplicably higher than 100 beats per min (bpm) at rest or higher than 90 bpm on average over 24 hours. The pathophysiology of IST is still not fully understood as the processes involved in pacemaking and its modulation are extremely complex (Baruscotti et al., 2016; Olshansky and Sullivan, 2019). Any participating mechanisms can affect the HR and result in this syndrome, including an intrinsic increase in sinoatrial node automaticity (Baruscotti et al., 2016), excess sympathetic tone and reduced cardiovagal tone (Nwazue et al., 2014), and circulating anti-beta-adrenergic receptor antibodies (Chiale et al., 2006), among others.

The hyperpolarization-activated cyclic nucleotide-gated (HCN) channel family comprises 4 members (HCN1-4) and have the peculiarity that they are activated by hyperpolarization rather than depolarization (Biel et al., 2009). These channels are expressed in the heart and in the central and peripheral nervous systems and exhibit a unique ion selectivity since they allow the passage of a depolarizing mixed Na^+^ and K^+^ current (Biel et al., 2009). In the heart, the current is known as pacemaker or funny current (I_f_) (because of its quite unusual biophysical profile) and has a key role in controlling the rhythmic activity in sinoatrial node cells (Biel et al., 2009; DiFrancesco, 2010; Hennis et al., 2022; Rivolta et al., 2020). Neuronal current is generally designated as hyperpolarization-activated current or I_h_ and contributes to controlling neuronal excitability. HCN channels contain a cyclic nucleotide-binding domain (CNBD) in the carboxyl terminus connected to the pore-forming S6 transmembrane segment via the C-linker (Lee and MacKinnon, 2017). cAMP or cGMP binding to the CNBD induces a rightward shift of the voltage dependence of HCN4 channel activation (DiFrancesco and Tortora, 1991; Wainger et al., 2001), thus increasing the current. Furthermore, all HCN channel isoforms contain an N-terminal domain (HCND) that, in HCN1 and HCN2 has been demonstrated to stabilize the closed pore in the setting of a depolarized voltage sensor and that physically interacts with the CNBD and the voltage-sensor domain (VSD) (Lee and MacKinnon, 2017; Porro et al., 2019). HCN family members differ in their sensitivity to cAMP, opening kinetics, and preferential tissue distribution (Biel et al., 2009). It is generally accepted that in all mammalian species studied so far, HCN4 was found to be the main HCN channel isoform in the sinoatrial node although HCN1 and HCN2 isoforms are also present (Baruscotti et al., 2016, 2005; Shi et al., 1999). However, there are data suggesting that HCN1 is the predominant isoform expressed in the human sinoatrial node (Li et al., 2021, 2015).

In a large Spanish family with IST, we identified a novel heterozygous mutation (p.V240M) within the HCND of HCN4 channels. Understanding the pathophysiology of IST is crucial to find a treatment for this debilitating condition. For its part, the gating properties of HCN4 channels and their role in cardiac pacemaking are currently not fully understood. Thus, we decided to clinically study all family members and functionally analyze the p.V240M HCN4 mutation through biochemical and electrophysiological approaches even at the single-channel level. Our results demonstrated that p.V240M is the first naturally occurring HCND gain-of-function mutation causing an increased inward current flow during diastole accounting for the faster-than-normal heart rate of all carriers, that was reversed with treatment with ivabradine, a selective HCN channel blocker (Baruscotti et al., 2005). Moreover, at the molecular level our findings demonstrate the critical importance of the HCND in the gating o Our results strengthen the importance of genetic and functional studies in families carrying mutations with uncertain significance since they allow the implementation of a personalized treatment f HCN4 channels. and help to the understanding of the mechanisms controlling human cardiac electrophysiological properties.

## RESULTS

### Clinical and genetic characteristics of the patients

The proband (III.3 in Figure 1A) is a female in her 30’s (31-35), with a mean 24-h HR of 106[17] bpm (Table 1 and Figure 1B). After a thorough evaluation, no reversible causes of sinus tachycardia were found (i.e., hyperthyroidism, anemia, diabetes, orthostatic hypotension, infections, and drug abuse), thus establishing the diagnosis of IST. The echocardiogram and cardiac magnetic resonance (CMR) showed mild systolic dysfunction [left ventricular ejection fraction (LVEF)=43%], and no gadolinium enhancement. No signs of left ventricular noncompaction or hypertrabeculation were found. Her resting HR and electrocardiographic (ECG, in at least three different recordings) and cardiac structural parameters are described in Table 2. Clinical evaluation of her relatives revealed that her father, two siblings, two sons, and three nephews also fulfilled diagnostic criteria of IST, with a mean resting HR of 113[7] bpm (Table 1 and Figure 1).

**Table 1.**
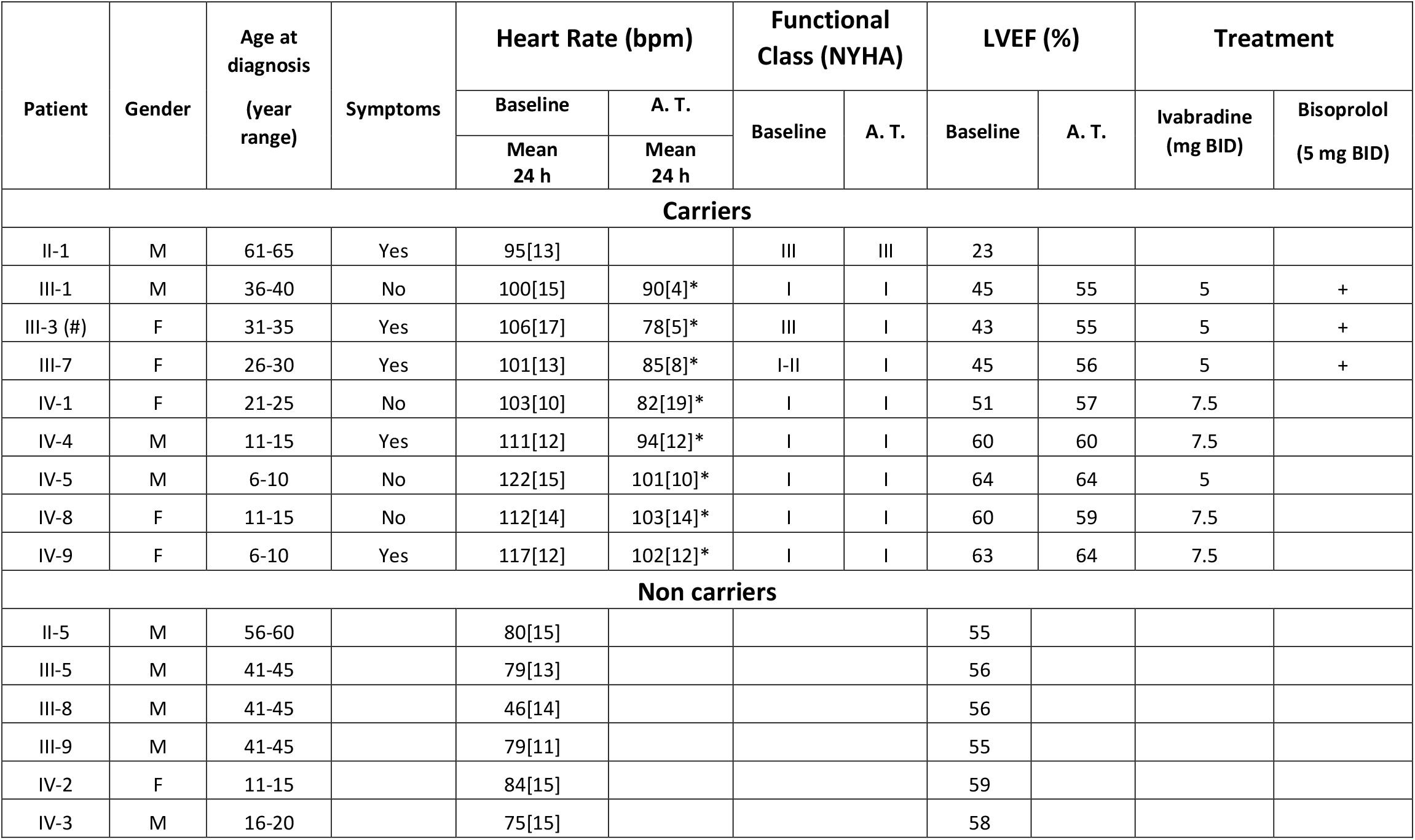
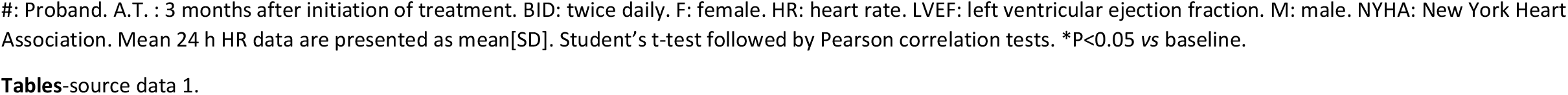
Clinical characteristics of the family members studied.

| Patient | Gender | Age at diagnosis<br>(year range) | Symptoms | Heart Rate (bpm) |  | Functional Class (NYHA) |  | LVEF (%) |  | Treatment |  |
| --- | --- | --- | --- | --- | --- | --- | --- | --- | --- | --- | --- |
|  |  |  |  | Baseline | A. T. | Baseline | A. T. | Baseline | A. T. | Ivabradine (mg BID) | Bisoprolol (5 mg BID) |
|  |  |  |  | Mean 24 h | Mean 24 h |  |  |  |  |  |  |
| Carriers |  |  |  |  |  |  |  |  |  |  |  |
| II-1 | M | 61-65 | Yes | 95[13] |  | III | III | 23 |  |  |  |
| III-1 | M | 36-40 | No | 100[15] | 90[4]* | I | I | 45 | 55 | 5 | + |
| III-3 (#) | F | 31-35 | Yes | 106[17] | 78[5]* | III | I | 43 | 55 | 5 | + |
| III-7 | F | 26-30 | Yes | 101[13] | 85[8]* | I-II | I | 45 | 56 | 5 | + |
| IV-1 | F | 21-25 | No | 103[10] | 82[19]* | I | I | 51 | 57 | 7.5 |  |
| IV-4 | M | 11-15 | Yes | 111[12] | 94[12]* | I | I | 60 | 60 | 7.5 |  |
| IV-5 | M | 6-10 | No | 122[15] | 101[10]* | I | I | 64 | 64 | 5 |  |
| IV-8 | F | 11-15 | No | 112[14] | 103[14]* | I | I | 60 | 59 | 7.5 |  |
| IV-9 | F | 6-10 | Yes | 117[12] | 102[12]* | I | I | 63 | 64 | 7.5 |  |
| Non carriers |  |  |  |  |  |  |  |  |  |  |  |
| II-5 | M | 56-60 |  | 80[15] |  |  |  | 55 |  |  |  |
| III-5 | M | 41-45 |  | 79[13] |  |  |  | 56 |  |  |  |
| III-8 | M | 41-45 |  | 46[14] |  |  |  | 56 |  |  |  |
| III-9 | M | 41-45 |  | 79[11] |  |  |  | 55 |  |  |  |
| IV-2 | F | 11-15 |  | 84[15] |  |  |  | 59 |  |  |  |
| IV-3 | M | 16-20 |  | 75[15] |  |  |  | 58 |  |  |  |

#: Proband. A.T. : 3 months after initiation of treatment. BID: twice daily. F: female. HR: heart rate. LVEF: left ventricular ejection fraction. M: male. NYHA: New York Heart Association. Mean 24 h HR data are presented as mean[SD]. Student's t-test followed by Pearson correlation tests. \*P<0.05 vs baseline.

**Table 2.** Resting ECG and cardiac structural parameters of all the family members studied.

| Patient | ECG parameters |  |  |  |  |  |  |  |  |  |  |  | Chamber characterisation |  |  |  |
| --- | --- | --- | --- | --- | --- | --- | --- | --- | --- | --- | --- | --- | --- | --- | --- | --- |
|  | Resting Heart Rate (bpm) |  | P (ms) |  | PQ (ms) |  | QRS (ms) |  | QT (ms) |  | QTc (ms) |  | LVEDD (mm) |  | LVESD (mm) |  |
|  | Baseline | A.T. | Baseline | A. T. | Baseline | A. T. | Baseline | A. T. | Baseline | A. T. | Baseline | A. T. | Baseline | A. T. | Baseline | A. T. |
| **CARRIERS** |  |  |  |  |  |  |  |  |  |  |  |  |  |  |  |  |
| **II-1** | 117 | - | 100 | - | 120 | - | 96 | - | 310 | - | 380 | - | 53 | - | 44 | - |
| **III-1** | 106 | 73 | 110 | 120 | 140 | 150 | 90 | 90 | 320 | 360 | 390 | 400 | 55 | 56 | 43 | 39 |
| **III-3 (#)** | 107 | 77 | 120 | 120 | 200 | 160 | 80 | 90 | 320 | 360 | 390 | 400 | 53 | 52 | 40 | 40 |
| **III-7** | 108 | 83 | 100 | 100 | 160 | 160 | 80 | 100 | 320 | 340 | 390 | 400 | 50 | 53 | 33 | 40 |
| **IV-1** | 102 | 73 | 100 | 100 | 160 | 160 | 80 | 80 | 340 | 360 | 400 | 400 | 46 | 44 | 36 | 31 |
| **IV-4** | 119 | 90 | 80 | 100 | 180 | 200 | 80 | 80 | 310 | 340 | 390 | 410 | 49 | 50 | 33 | 34 |
| **IV-5** | 123 | 100 | 80 | 80 | 140 | 160 | 80 | 90 | 300 | 340 | 380 | 400 | 41 | 40 | 27 | 25 |
| **IV-8** | 123 | 87 | 80 | 100 | 120 | 140 | 80 | 80 | 300 | 340 | 380 | 420 | 45 | 46 | 31 | 30 |
| **IV-9** | 114 | 94 | 120 | 120 | 160 | 166 | 90 | 100 | 320 | 340 | 400 | 420 | 41 | 42 | 26 | 27 |
| **NON CARRIERS** |  |  |  |  |  |  |  |  |  |  |  |  |  |  |  |  |
| **II-5** | 61 | - | 100 | - | 140 | - | 90 | - | 380 | - | 380 | - | 48 | - | 34 | - |
| **III-5** | 75 | - | 100 | - | 180 | - | 80 | - | 340 | - | 380 | - | 50 | - | 34 | - |
| **III-8** | 81 | - | 100 | - | 160 | - | 90 | - | 360 | - | 400 | - | 52 | - | 40 | - |
| **III-9** | 80 | - | 80 | - | 140 | - | 80 | - | 340 | - | 410 | - | 50 | - | 34 | - |
| **IV-2** | 82 | - | 80 | - | 120 | - | 80 | - | 360 | - | 410 | - | 43 | - | 30 | - |
| **IV-3** | 53 | - | 100 | - | 160 | - | 90 | - | 400 | - | 420 | - | 50,2 | - | 34,8 | - |

A.T.: 3 months after treatment. bpm: beats per minute. ECG: electrocardiogram. F: female. HR: heart rate. LVEDD: left ventricular end-diastolic diameter. LVEF: left ventricular ejection fraction. LVESD: left ventricular end-systolic diameter. QT values were corrected using the Bazett's and Fredericia's formulas in subjects with HR <100 and ≥100 bpm, respectively. Statistical comparison between baseline measurements and data after treatment was analyzed by paired Student t-tests.
**Tables-**source data 1.

**Figure 1.**
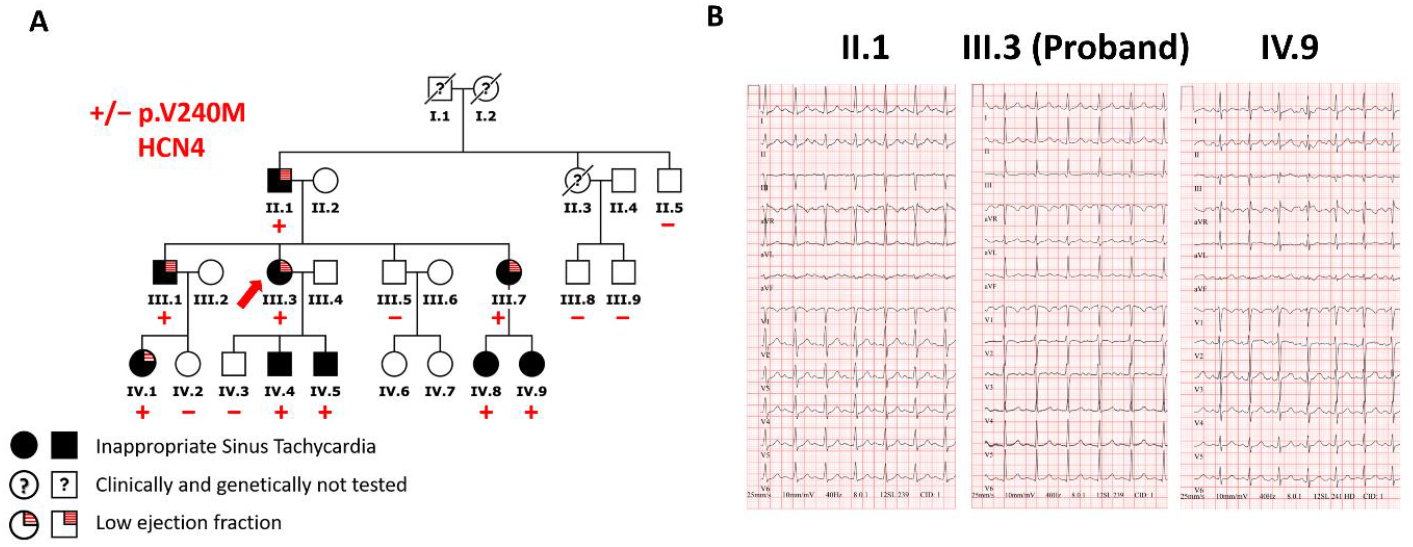
A family with inappropriate sinus tachycardia. (**A**) Pedigree of the IST proband’s family. The arrow indicates the proband (III.3), circles and squares represent females and males, respectively. (**B**) Twelve-lead ECGs of II.1, III.3, and IV.9 family members (paper speed 25 mm/s).

Six more relatives were also clinically studied and none of them was diagnosed with IST (Table 1). No significant electrocardiographic or structural findings were detected in any of the IST patients (Table 2). However, all adults with IST (5 out of 9) had reduced LVEF without gadolinium enhancement, whereas children did not have cardiac dysfunction (Table 1 and Figure 1). As expected, the HR of children was higher than that of adults (mean 24-h HR: 115.2[5.3] *vs*. 102.7[2.1] bpm, P<0.05, n≥4). Patients IV.4 and IV.8 were diagnosed with fetal tachycardia with HR at 160-170 bpm at birth (>98 percentile). The proband’s aunt (II.3) had died after heart transplant due to a congenital heart defect.

These data suggested that in this family there was a genetic component responsible for the IST. Next-generation sequencing (Table Supplement File 1) of the proband identified the pathogenic (according to the guidelines for the interpretation of variants, Table 3 and Methods) heterozygous 15,73659894,C,T (NM_005477.3:c.718G>A) variant in *HCN4* gene encoding p.V240M HCN4 (Figure 2A). The mutated residue is highly conserved among different species and lays in the HCND of the protein (Figure 2 B-D).

**Table 3.**
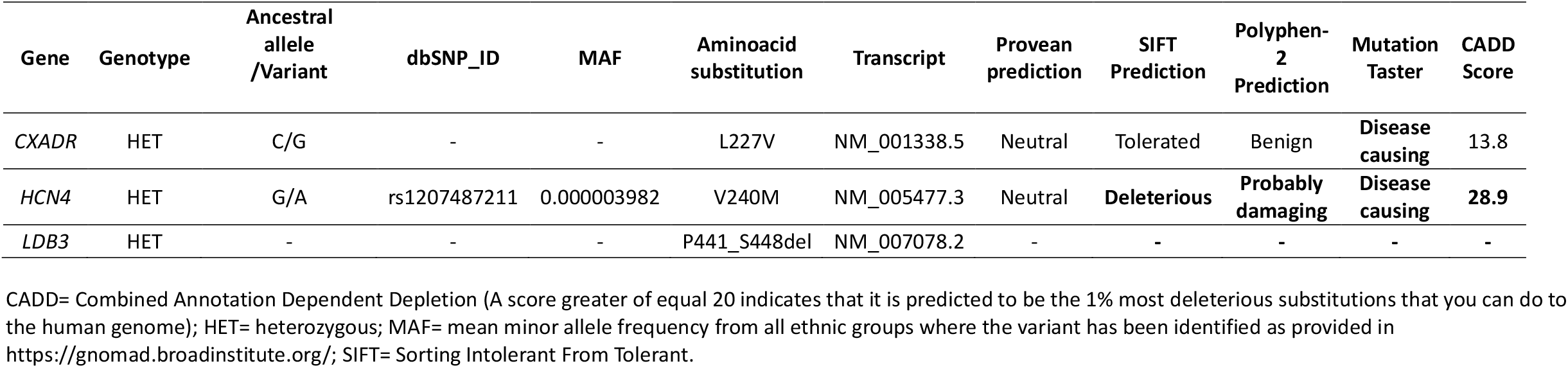
Summary of all nonsynonymous variants identified in the proband.

| Gene | Genotype | Ancestral allele /Variant | dbSNP_ID | MAF | Aminoacid substitution | Transcript | Provean prediction | SIFT Prediction | Polyphen-2 Prediction | Mutation Taster | CADD Score |
| --- | --- | --- | --- | --- | --- | --- | --- | --- | --- | --- | --- |
| *CXADR* | HET | C/G | - | - | L227V | NM_001338.5 | Neutral | Tolerated | Benign | Disease causing | 13.8 |
| *HCN4* | HET | G/A | rs1207487211 | 0.000003982 | V240M | NM_005477.3 | Neutral | Deleterious | Probably damaging | Disease causing | 28.9 |
| *LDB3* | HET | - | - | - | P441_S448del | NM_007078.2 | - | - | - | - | - |

CADD= Combined Annotation Dependent Depletion (A score greater of equal 20 indicates that it is predicted to be the 1% most deleterious substitutions that you can do to the human genome); HET= heterozygous; MAF= mean minor allele frequency from all ethnic groups where the variant has been identified as provided in https://gnomad.broadinstitute.org/; SIFT= Sorting Intolerant From Tolerant.

**Figure 2.**
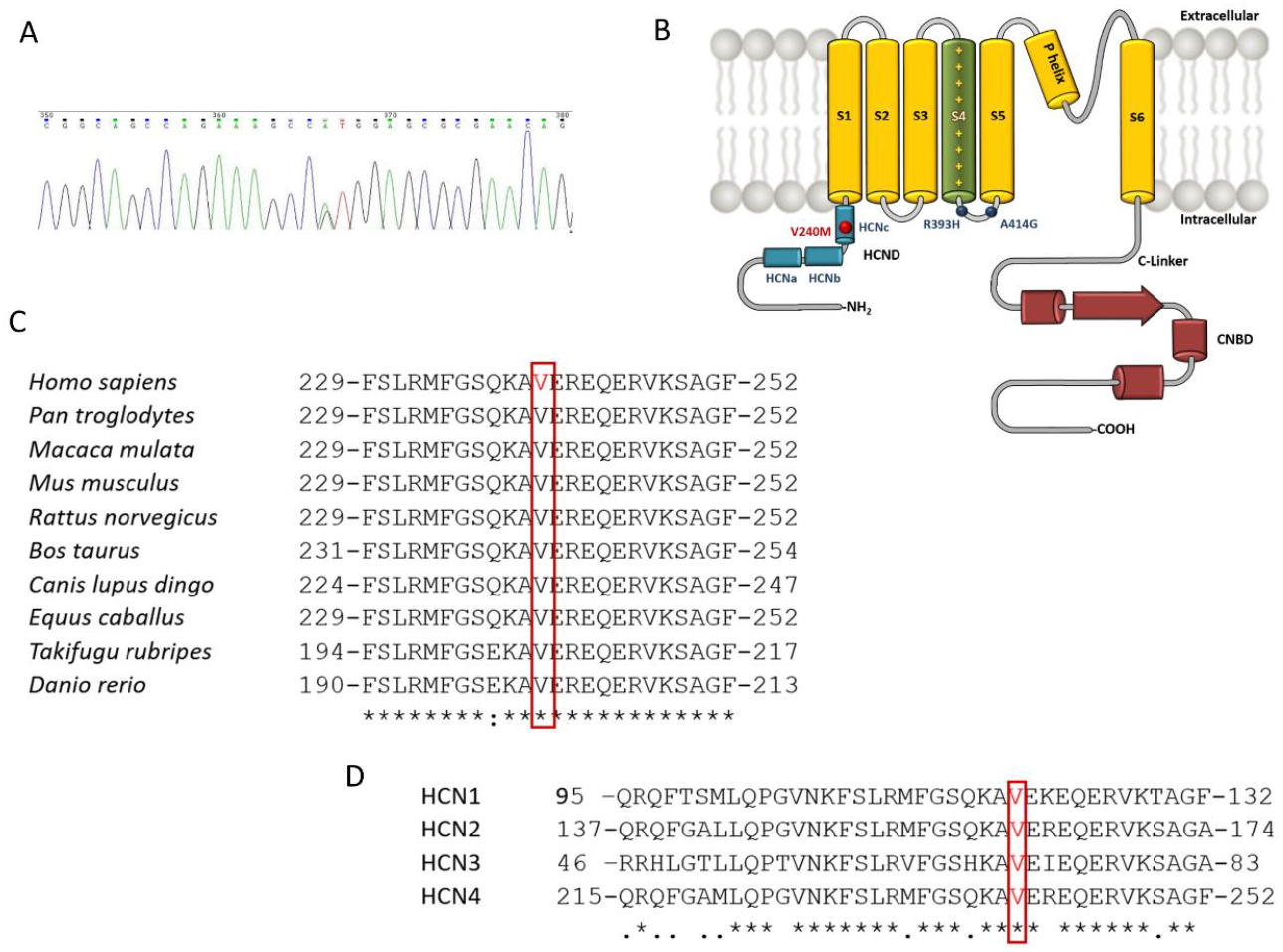
The V240 residue is highly conserved and lays in the HCND of the protein. (**A**) DNA sequence chromatogram depicting the heterozygous *HCN4* variation (15,73659894,C,T; c.718G>A) in the proband. (**B**) Schematic diagram of human HCN4 channel highlighting some key domains and the approximate location of the p.V240M variant. (**C** and **D**) Sequence alignment of the region surrounding the V240 residue in HCN4 of several species (**C**) or in the four channels of the HCN family (**D**). The boxes in panels C and D highlight the conservation of this residue.

The p.V240M variant has been previously annotated with an allele frequency of 0.000003982 (https://gnomad.broadinstitute.org, accession date: 11/21/2022) since it was detected on a single allele in a south Asian subject. Cascade screening demonstrated the genotype-phenotype co-segregation of the *HCN4* variant (Figure 1A). All carriers except II.1, who refused any treatment, were initially treated with ivabradine alone (7.5 mg BID for adults and 5 mg BID for the youngest carrier) with a significant response in terms of mean 24-h HR decrease (94[7] vs. 109[7.9], P<0.05) and improvement in symptoms and quality of life (Table 1). Three adult patients had their ivabradine dose reduced (5 mg BID) since they experienced mild weakness and dizziness, which are frequently associated with the use of this drug. Bisoprolol (5 mg BID) was added to them to achieve a better HR control (from 91[1.5] before to 84[5] after bisoprolol).

### Macroscopic current analysis

To test whether the p.V240M HCN4 mutation underlies the IST in this family we functionally analyzed this variant. Figure 3A shows macroscopic HCN4 current (I_HCN4_) traces generated in three different CHO cells transfected with WT, p.V240M, and the combination of WT and p.V240M HCN4 channels (0.5:0.5 ratio). I_HCN4_ comprises two components (inset in Figure 3A): the minor instantaneous (I_INS_) and the major slowly developing steady-state current (I_SS_), whose amplitude progressively increased at more negative potentials (Biel et al., 2009; Mistrík et al., 2006).

**Figure 3.**
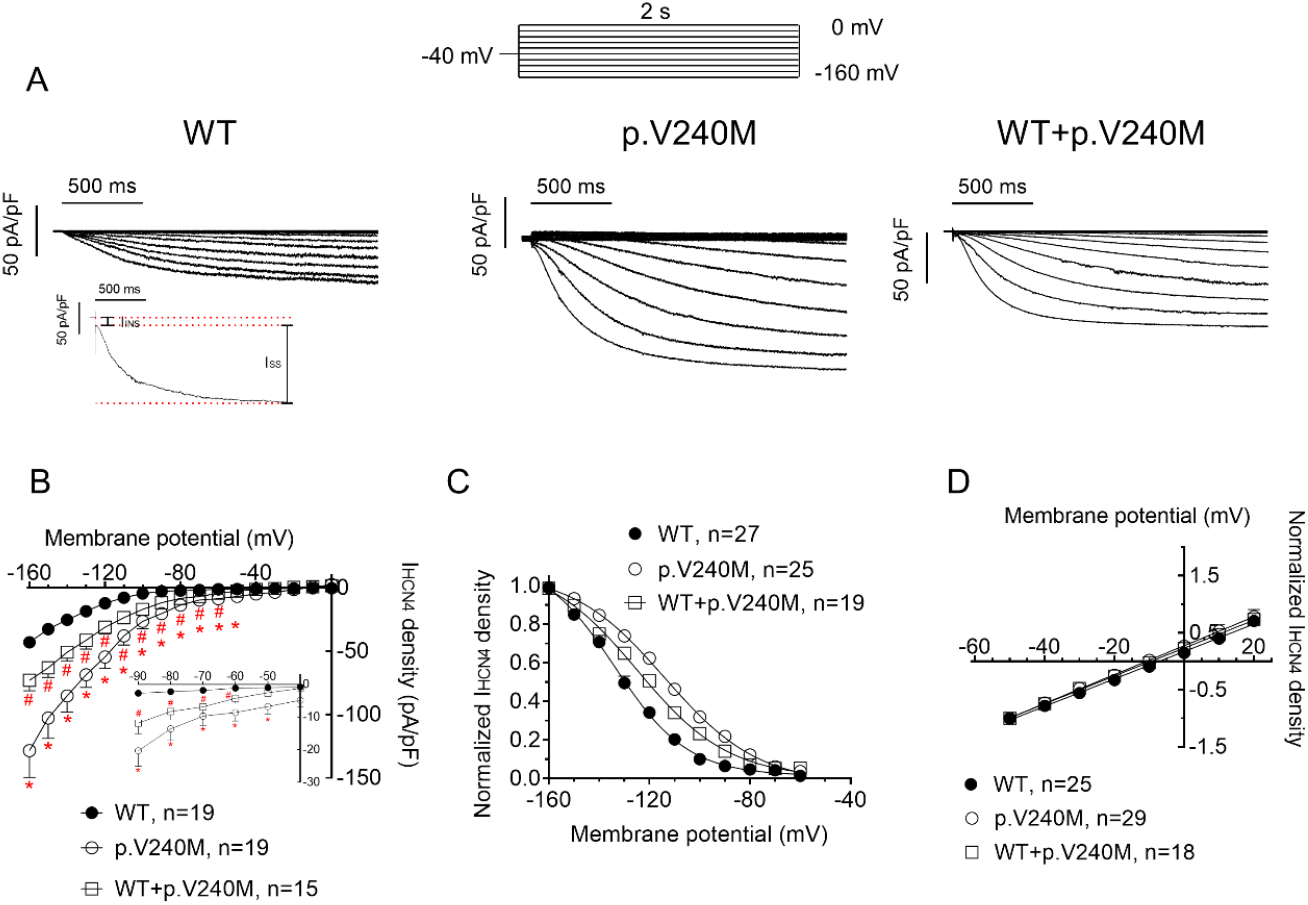
Macroscopic currents generated by WT and p.V240M HCN4 channels. (**A** and **B**) I_HCN4_ traces (**A**) and density–voltage relationships (**B**) generated in CHO cells transiently expressing WT, p.V240M, and WT+p.V240M HCN4 channels by applying the protocol depicted at the top. The inset in A shows the minor instantaneous (I_INS_) and the major slowly developing steady-state HCN4 current (I_SS_) and the inset in B, shows the data at potentials positive to -90 mV in an expanded scale. (**C**) Normalized tail current densities generated by WT, p.V240M, and WT+p.V240M channels by applying 1-second pulses to −140 mV were plotted against the membrane potential of the test pulse. Continuous lines represent the Boltzmann fit to the data. (**D**) Fully activated I_HCN4_ density–voltage relationships generated by WT, p.V240M, and WT+p.V240M channels. Continuous lines represent the linear regression to the data. In **B**-**D**, each point represents the mean±SEM of ≥15 experiments/cells (biological replicates) from ≥3 dishes. In B, *P<0.05 vs. HCN4 WT. #P<0.05 vs. HCN4 WT and P<0.05 vs. p.V240M. Two-way ANOVA and Multiple t-test. **Figure 3-**source data 1 of panels B, C, and D.

I_HCN4_ generated by p.V240M channels (measured at the end of the pulses) was significantly greater than that generated by WT channels (Figure 3A and 3B) in a wide range of membrane potentials that includes physiological membrane potentials (−1.2±0.2 *vs*. -8.6±2.6 pA/pF at -60 mV; P<0.05, n=19) (Table 4). Interestingly, densities of I_HCN4_ generated by the cotransfection of WT and p.V240M channels were also significantly greater than those of WT channels (Figure 3A and 3B and Table 4). Figure 3C shows that the p.V240M variant either alone or in combination with WT channels, significantly and markedly shifted the midpoint (Vh) of the activation curve to more depolarized membrane potentials as assessed by Boltzmann fits (Table 4).

**Table 4.**
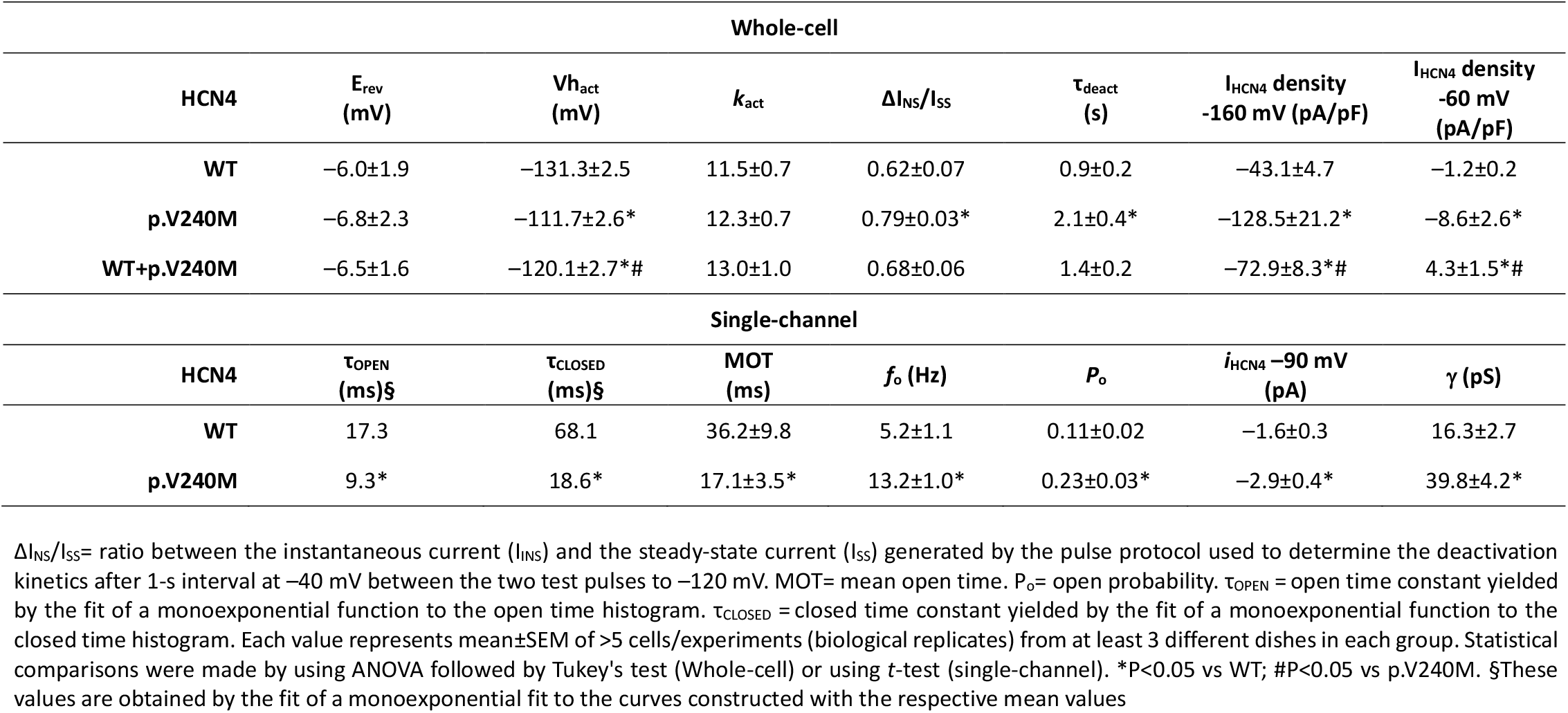
Effects of p.V240M variant on the time- and voltage-dependent properties of macroscopic and unitary HCN4 currents

| Whole-cell |  |  |  |  |  |  |  |
| --- | --- | --- | --- | --- | --- | --- | --- |
| HCN4 | $E_{rev}$<br>(mV) | $V_{h_{act}}$<br>(mV) | $k_{act}$ | $\Delta I_{NS}/I_{SS}$ | $\tau_{deact}$<br>(s) | $I_{HCN4}$ density<br>-160 mV (pA/pF) | $I_{HCN4}$ density<br>-60 mV<br>(pA/pF) |
| WT | -6.0±1.9 | -131.3±2.5 | 11.5±0.7 | 0.62±0.07 | 0.9±0.2 | -43.1±4.7 | -1.2±0.2 |
| p.V240M | -6.8±2.3 | -111.7±2.6* | 12.3±0.7 | 0.79±0.03* | 2.1±0.4* | -128.5±21.2* | -8.6±2.6* |
| WT+p.V240M | -6.5±1.6 | -120.1±2.7*# | 13.0±1.0 | 0.68±0.06 | 1.4±0.2 | -72.9±8.3*# | 4.3±1.5*# |
| Single-channel |  |  |  |  |  |  |  |
| HCN4 | $\tau_{OPEN}$<br>(ms)§ | $\tau_{CLOSED}$<br>(ms)§ | MOT<br>(ms) | $f_o$ (Hz) | $P_o$ | $i_{HCN4}$ -90 mV<br>(pA) | $\gamma$ (pS) |
| WT | 17.3 | 68.1 | 36.2±9.8 | 5.2±1.1 | 0.11±0.02 | -1.6±0.3 | 16.3±2.7 |
| p.V240M | 9.3* | 18.6* | 17.1±3.5* | 13.2±1.0* | 0.23±0.03* | -2.9±0.4* | 39.8±4.2* |

$\Delta I_{NS}/I_{SS}$ = ratio between the instantaneous current ( $I_{INS}$ ) and the steady-state current ( $I_{SS}$ ) generated by the pulse protocol used to determine the deactivation kinetics after 1-s interval at -40 mV between the two test pulses to -120 mV. MOT= mean open time. $P_o$ = open probability. $\tau_{OPEN}$ = open time constant yielded by the fit of a monoexponential function to the open time histogram. $\tau_{CLOSED}$ = closed time constant yielded by the fit of a monoexponential function to the closed time histogram. Each value represents mean±SEM of >5 cells/experiments (biological replicates) from at least 3 different dishes in each group. Statistical comparisons were made by using ANOVA followed by Tukey's test (Whole-cell) or using *t*-test (single-channel). \* $P$ <0.05 vs WT; # $P$ <0.05 vs p.V240M. §These values are obtained by the fit of a monoexponential fit to the curves constructed with the respective mean values

Activation time constant (τ_act_) at -160 mV averaged 509±40, 354±42, and 438±40 ms for I_HCN4_ generated by WT, p.V240M, or WT+p.V240M HCN4 channels, respectively (Figure 4). Thus, the p.V240M mutation either alone or in combination with WT channels, significantly accelerated the activation of I_HCN4_ (P=0.026, n≥17).

**Figure 4.**
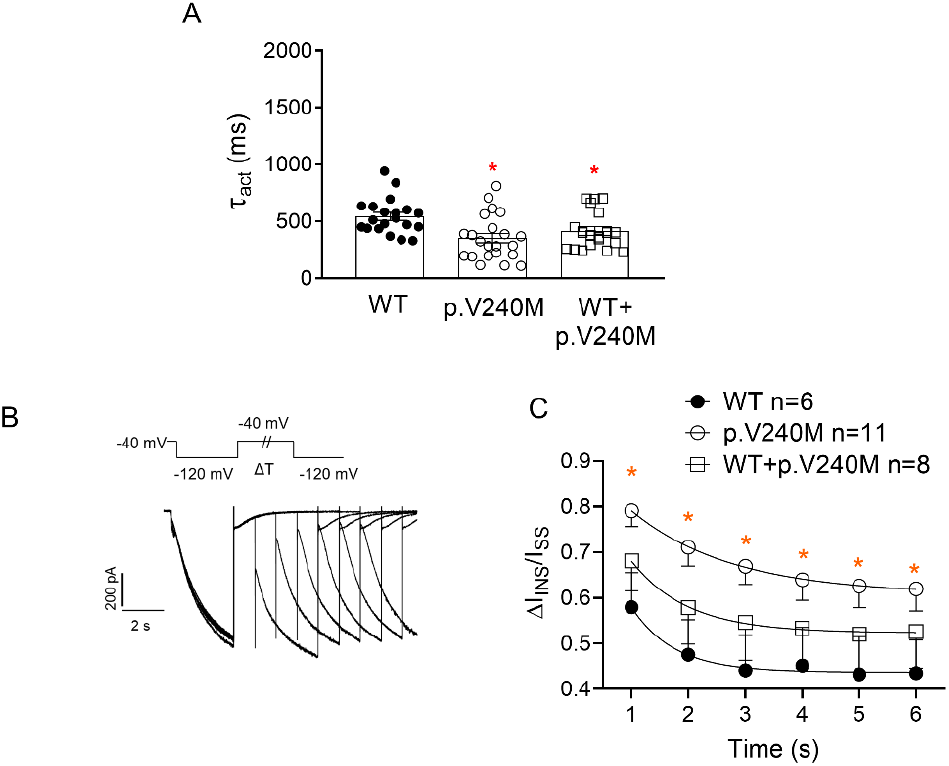
p.V240M HCN4 modifies the time dependence of I_HCN4_ activation. (**A**) Mean time constant of activation (τ_act_) yielded by the fit of a monoexponential function to the activating phase of HCN4 current (I_HCN4_) traces elicited by pulses to -160 mV in cells expressing WT, p.V240M or WT+p.V240M channels. Each bar represents the mean±SEM of n experiments and each dot represents one experiment. (**B**) I_HCN4_ traces recorded by applying the protocol shown at the top to measure deactivation kinetics consisting of two 3-s pulses from –40 to –120 mV that were applied at increasing coupling intervals (1-6 s). For each coupling interval, we measured the instantaneous current (ΔI_INS_) and the steady-state current (I_SS_) elicited at the beginning and at the end, respectively, of the second pulse to –120 mV. (**C**) Mean ΔI_INS_/I_SS_ measured at each coupling interval for I_HCN4_ recorded in cells expressing WT, p.V240M or WT+p.V240M channels. A monoexponential function was fitted to the data to obtain the time constant of deactivation (τ_deact_). Each point was the mean±SEM of ≥7 experiments (biological replicates). In A and C, *P<0.05 vs WT. Two-way ANOVA and multiple t-test. An F-test was performed to determine the statistically significance of the differences between activation time constants obtained in E. **Figure 4**-source data 1 of panels A and C.

Channel deactivation properties were analyzed using a double pulse protocol (Schweizer et al., 2010). Figure 4C shows that deactivation of p.V240M channels alone, but not in combination with WT, was significantly slower than that of WT channels (P<0.05, n≥7) and, as a consequence, the ratio of p.V240M channels that remained activated is greater for all the interpulse intervals (Table 4).

### Effects of cAMP on p.V240M HCN4 channels

Interestingly, effects produced by the p.V240M variant are quite similar to those produced by cAMP (DiFrancesco and Tortora, 1991) and thus, we hypothesized that the mutation increases I_HCN4_ by augmenting HCN4 channel affinity for cAMP. Since HCN4 activation is not affected by cAMP when channels are expressed in CHO cells (Peters et al., 2020), we tested our hypothesis in HEK293 cells that were exposed to a saturating cAMP concentration (10 μM) in the whole-cell pipette solution.

As expected, cAMP significantly shifted the activation curve of WT HCN4 channels toward depolarizing potentials (Figure 5A). In HEK293 cells the Vh of p.V240M HCN4 channel activation was also significantly more depolarized than that of WT channels (−111.3±3.8 mV, n≥8, P<0.05). Importantly, voltage-dependent activation of p.V240M channels was significantly shifted in the presence of cAMP (−97.1±5.2 mV, n≥6, P<0.05) (Figure 5B).

**Figure 5.**
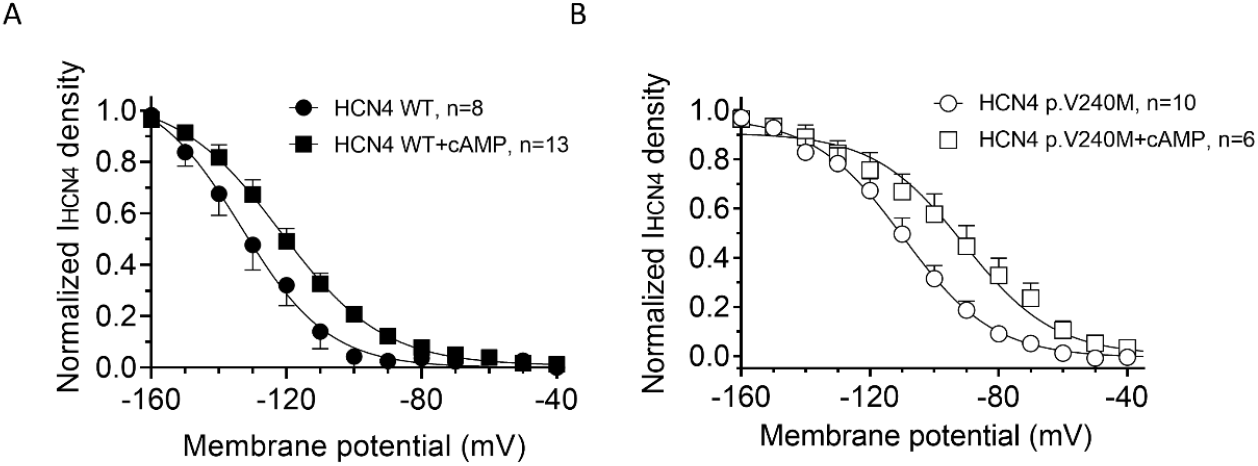
Effects of cAMP on the voltage dependence of I_HCN4_ activation. (**A** and **B**) Voltage dependence of I_HCN4_ generated by WT (**A**) and p.V240M (**B**) channels expressed in HEK293 cells in the presence or absence of 10 μM cAMP. Tail current densities generated by applying 1-s pulses to −140 mV were normalized and plotted against the membrane potential of the test pulse. Continuous lines represent the Boltzmann fit to the data. Each point represents the mean±SEM of ≥6 experiments/cells (biological replicates) from ≥3 dishes. **Figure 5**-source data 1 of panels A and B.

### Effects of the p.V240M HCN4 mutation on the membrane expression

To analyze whether an increase of the expression of p.V240M HCN4 channels at the plasma membrane accounted for the I_HCN4_ increase, we conducted a cell surface biotinylation assay (Figure 6) measuring the levels of expression of total HCN4 protein (inputs) and membrane fraction (biotinylated) by Western blot (WB) (Figure 6A). Figure 6B shows the densitometry of the relative surface expression of WT and p.V240M HCN4 subunits and demonstrates that there were not significant differences in the expression of mutated and WT channels (P>0.05, n=5).

**Figure 6.**
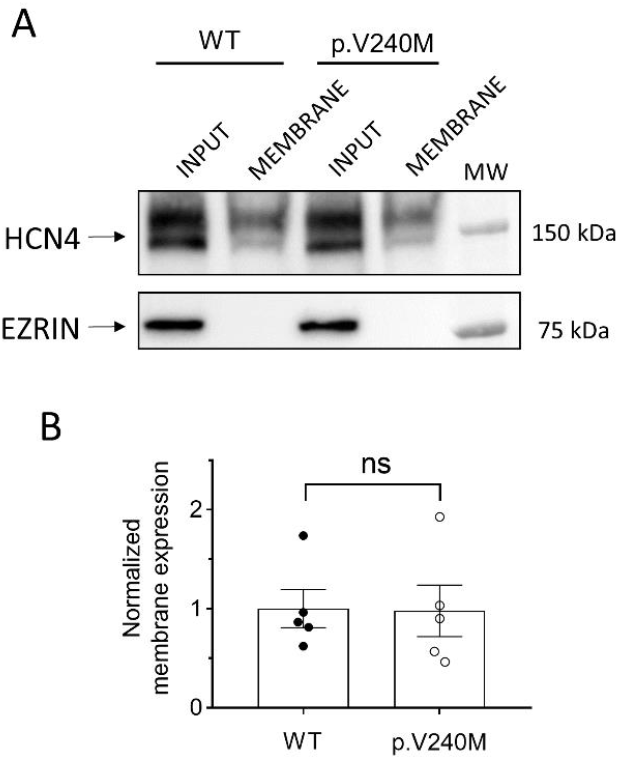
Membrane expression of WT and p.V240M HCN4 channels. Representative WB images (**A**) and densitometric analyses (**B**) of biotinylation assays showing the total (input) or surface (membrane) expression of HCN4 in cells expressing WT or p.V240M channels. The cytosolic protein ezrin was used as a negative control. In **B**, each dot represents 1 experiment and each bar the mean±SEM of n experiments (biological replicates). Unpaired Student t-test. **Figure 6**-source data 1 of panel A. **Figure 6**-source data 2 of panel B.

### Single-channel properties of p.V240M HCN4 channels

Figure 7 shows representative single-channel traces recorded in CHO cells at -90 mV in the absence (control, top panels) or the presence (bottom panels) of ivabradine (5 µM), a selective I_f_ inhibitor (Baruscotti et al., 2005), using the cell-attached configuration.

**Figure 7.**
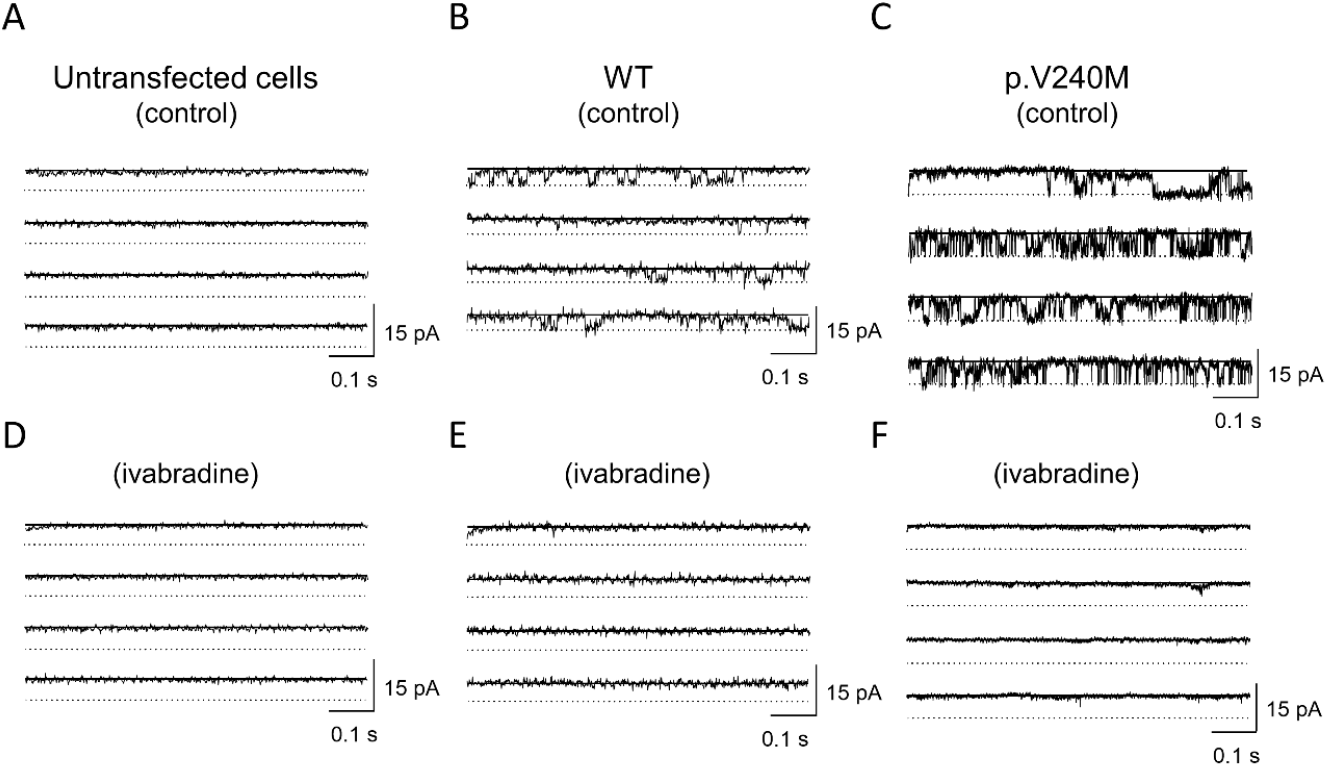
Single-channel recordings generated by WT and p.V240M HCN4 channels. Single channel recordings obtained by applying 3-s pulses from 0 to -90 mV in cells expressing or not (**A**) WT (**B**) or p.V240M (**C**) HCN4 channels in the absence and presence (**D-F**) of ivabradine 5 μM.

In all experiments we confirmed that ivabradine perfusion completely inhibited the unitary HCN4 current (*i*_HCN4_). Importantly, the p.V240M mutation did not modify the HCN4 channel sensitivity to ivabradine (Figure 8). No single-channel recordings were obtained in non-transfected cells since they do not express endogenous HCN4 channels (Figure 7A and 7D).

**Figure 8.**
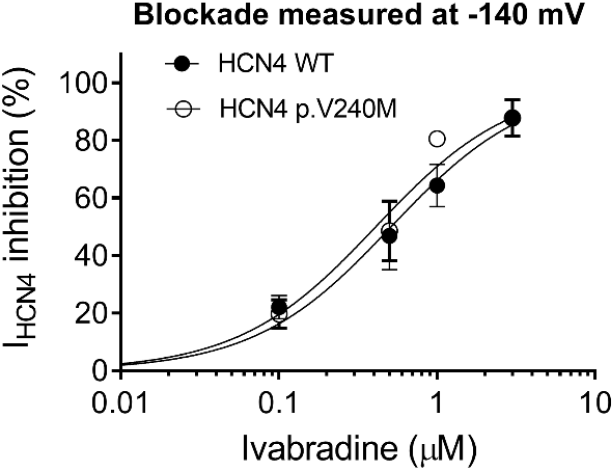
p.V240M does not modify HCN4 channel sensitivity to ivabradine. Concentration-response curves for ivabradine-induced inhibition of the I_HCN4_ recorded at –140 mV in cells expressing WT or p.V240M HCN4 channels. Solid lines represent the fit of a Hill equation to the data; the nH was fixed to unity, and bottom and top values to 0 and 100%, respectively. Each point was the mean±SEM of at least 3 experiments/cells (biological replicates). Statistical comparison between both curves was analyzed by an F-test (P=0.51). **Figure 8**-source data 1.

*i*_HCN4_ generated by WT channels (Figure 7B) was characterized by spontaneous openings followed by periods of channel closure yielding mean *f*_o_ and *P*_o_ values of 5.2±1.1 Hz and 0.11±0.02 (n=5), respectively (Figure 9A and 9B and Table 4).

**Figure 9.**
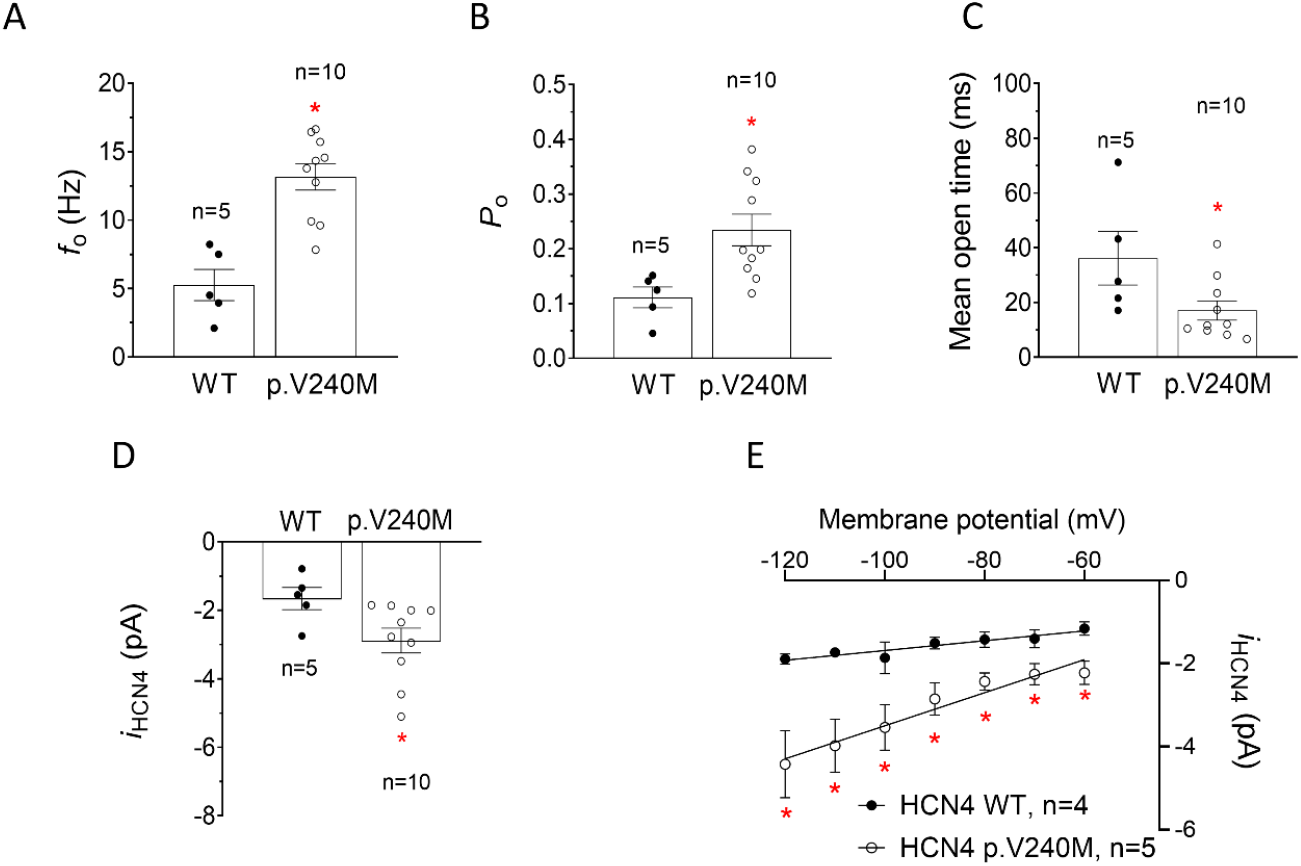
Gating properties of WT and p.V240M HCN4 channels. Mean *f*_o_ (**A**), *P*_o_ (**B**) and open time (**C**) for *i*_HCN4_ recorded in cells expressing WT or p.V240M HCN4 channels. (**D**) *i*_HCN4_ amplitude generated by WT and p.V240M channels after applying pulses to -90 mV. (**E**) *i*_HCN4_-voltage relationships generated by WT and p.V240M channels. In **A-D** each dot represents 1 experiment/cell and each bar the mean±SEM of n experiments/cells (biological replicates) as indicated in the figure. In **E** each point represents the mean±SEM of ≥4 experiments/cells (biological replicates). *P<0.05 vs. WT. Unpaired Student t-test. An F-test was performed to determine the statistically significance of the differences between conductance values. **Figure 9**-source data 1 of panels A-E.

Conversely, *i*_HCN4_ generated by p.V240M channels was characterized by openings in bursts (Figure 7C). Accordingly, both *f*_o_ and *P*_o_ significantly increased, while the mean open time significantly decreased compared to those of WT channels (P<0.05, n≥5) (Figure 9A-9C and Table 4). Moreover, *i*_HCN4_ amplitude generated by p.V240M channels at -90 mV (Figure 9D) and at all membrane potentials tested was significantly greater than that of WT channels (Figure 9E, P<0.05, n=5). Figure 9E shows that single-channel conductance (γ) derived from the linear regression of the current-voltage relationships of p.V240M channels was double than that of WT channels (Table 4). Importantly, γ here obtained for WT channels (16.3±2.7 pS) is in agreement with that previously reported for HCN4 channels (Brandt et al., 2009; Michels et al., 2005).

Gating kinetics was characterized by means of dwell-time histograms (Methods). Both close and open dwell-time histograms were fitted by monoexponential functions (Figure 10) yielding τ_CLOSED_ and τ_OPEN_ of 68.1 and 17.3 ms, respectively for WT channels. The mutation significantly reduced both the τ_CLOSED_ and the τ_OPEN_ (P<0.05) (Table 4 and Figure 10).

**Figure 10.**
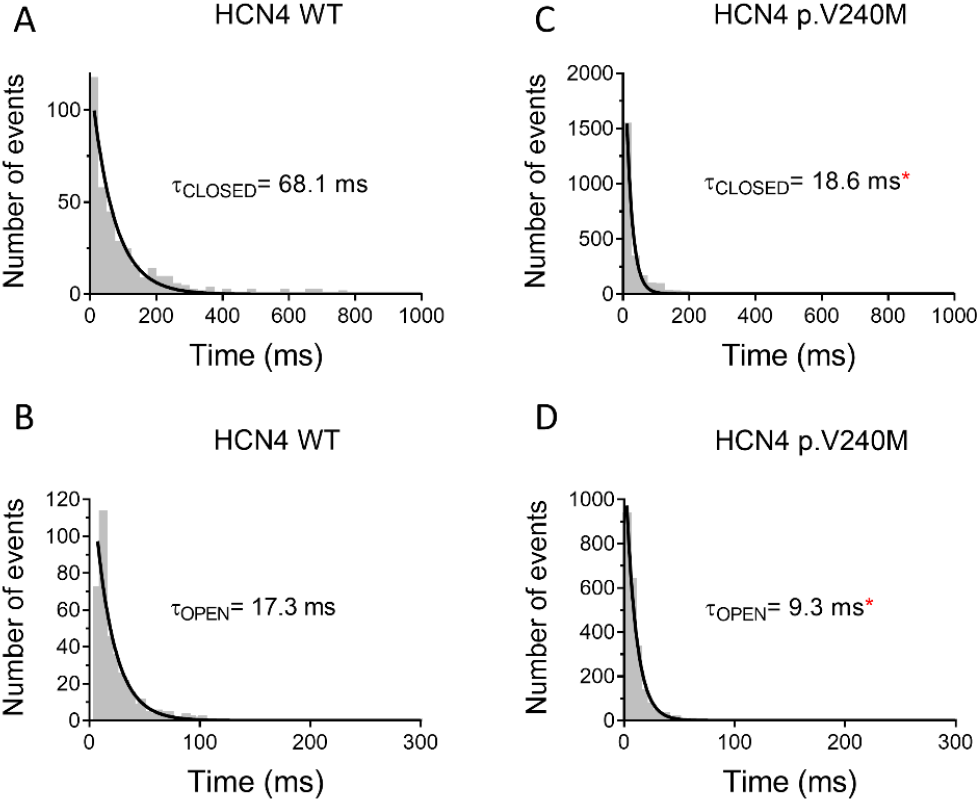
p.V240M modifies single channel gating kinetics. **(A-D)** Closed- (A and C) and open (B and D) dwell-time histograms for currents recorded in cells expressing HCN4 WT (A and B) or HCN4 p.V240M (C and D) (bin width= 25 ms). Continuous lines represent the fit of a monoexponential function to the data, which yielded the indicated closed (τ_CLOSED_) and open (τ_OPEN_) time constants. Histograms were obtained by pooling data from 5 (WT) and 10 (p.V240M) experiments (biological replicates). *P<0.05 vs WT (F-test). **Figure 10**-source data 1 of panels A-D.

### Mathematical model of human sinoatrial action potentials

Figure 11A shows human sinoatrial APs obtained when running the Fabbri-Fantini-Wilders-Severi mathematical model (Fabbri et al., 2017) considering the I_f_ generated by WT HCN4 channels. AP characteristics are described in Table 5. Spontaneous firing frequency of the AP averaged 73.8 bpm and it was almost doubled (146.3 bpm) when APs were modeled considering the I_f_ generated by p.V240M channels (Figure 11A). The acceleration of the pacemaker frequency is consequence of the huge increase in the I_f_ density produced by the mutation (Figure 11B). Interestingly, I_f_ currents generated when considering the heterozygous presence of p.V240M channels are also greater than those generated by WT channels and the firing frequency (108 bpm) is close to that of the carriers of the mutation.

**Table 5.** Characteristics of the AP generated by a mathematical model of the human sinoatrial node action potential

| Parameter | WT | p.V240M | WT+p.V240M |
| --- | --- | --- | --- |
| Cycle Length (ms) | 814 | 410 | 555 |
| Frequency (Hz/bpm) | 1.23/73.8 | 2.44/146.3 | 1.80/108.1 |
| Diastolic depolarization rate (mV/s) | 48.1 | 126.7 | 76.7 |
| Mean Diastolic Potential (mV) | -58.9 | -57.5 | -58 |
| Action potential amplitude (mV) | 85.3 | 81.5 | 84.0 |
| Overshoot (mV) | 26.4 | 24.0 | 26.0 |
| APD<sub>20</sub> (ms) | 98.5 | 85.0 | 93.2 |
| APD<sub>50</sub> (ms) | 136 | 122.9 | 129.6 |
| APD<sub>90</sub> (ms) | 161.5 | 148.7 | 155.0 |

APD<sub>20</sub>, APD<sub>50</sub>, APD<sub>90</sub>= action potential duration measured at 20%, 50% and 90% of repolarization; bpm= beats per minute.

**Figure 11.**
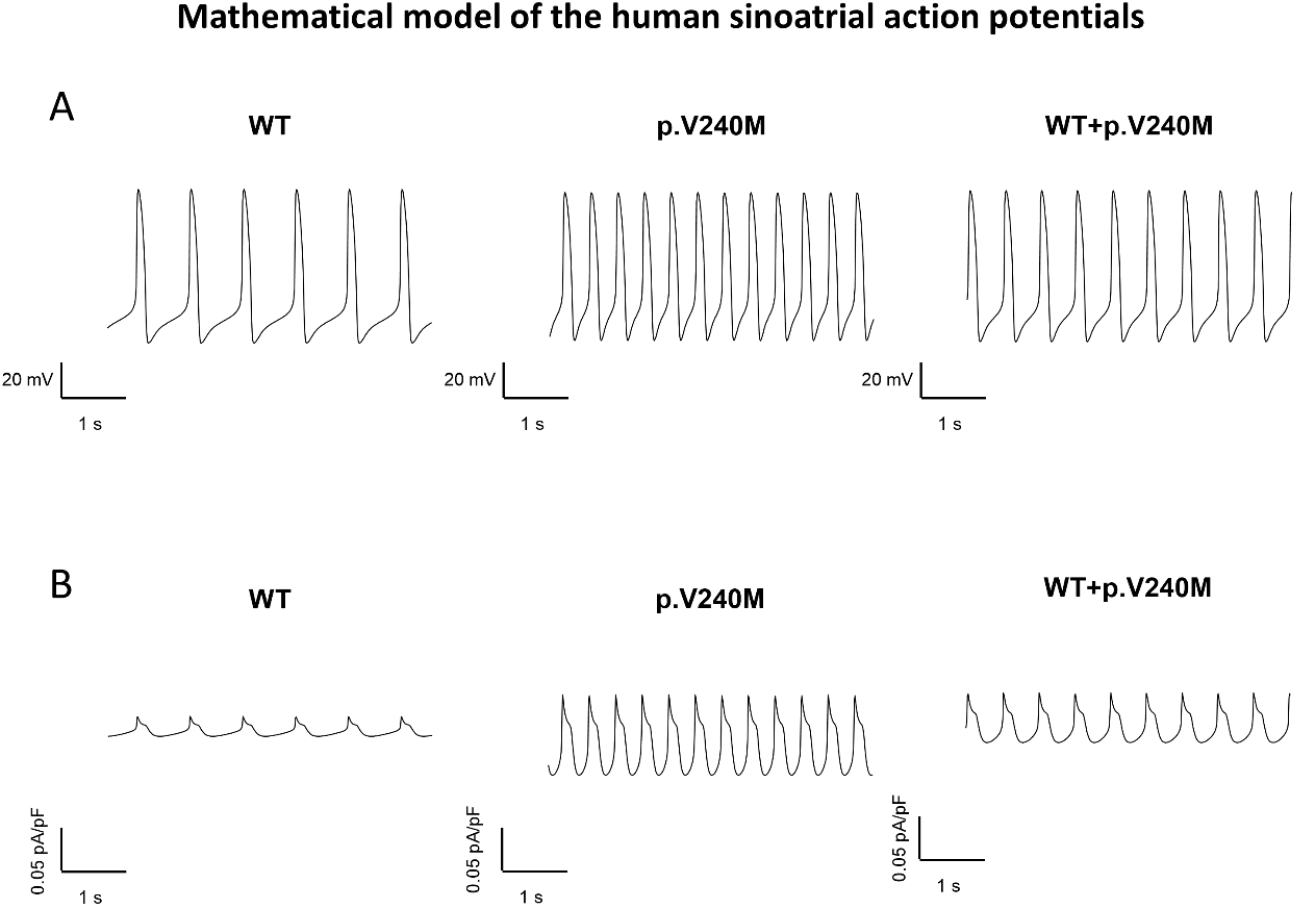
Mathematical model of human sinoatrial node cell action potential. (**A** and **B**). APs (**A**) and I_f_ (**B**) obtained when running the Fabbri-Fantini-Wilders-Severi model under basal conditions (WT; left), or after introducing the experimentally measured I_HCN4_ changes induced by p.V240M alone (center) or together with WT (right).

## DISCUSSION

In this study we identified several members of a family diagnosed with IST carrying a novel gain-of-function *HCN4* variant. The results of our functional analysis demonstrated that p.V240M variant markedly increases the *f*_o_ and the *P*_o_ and depolarizes the voltage dependence of HCN4 channels activation. Thus, the mutation increases I_HCN4_ by altering the gating without modifying the channel sensitivity for cAMP and ivabradine or the membrane expression. All these effects account for the huge increase of I_HCN4_, which, in turn, explains the IST of the affected family members who were treated with ivabradine, which reversed IST and tachycardia-induced cardiomyopathy.

The p.V240M mutation is, to the best of our knowledge, the second HCN4 gain-of-function mutation described in the literature. The first one was the p.R524Q HCN4 variant, that increases the channel affinity for cAMP and was described in an Italian family with symptomatic sinus tachyarrhythmia (Baruscotti et al., 2017). Conversely, many loss-of-function heterozygous mutations in the *HCN4* gene have been associated with sinus bradycardia that can be accompanied, or not, with atrial fibrillation, atrioventricular block, structural diseases such as noncompaction cardiomyopathy, and even QT prolongation (Alonso-Fernández-Gatta et al., 2021; Cambon-Viala et al., 2021; Macri et al., 2014; Milanesi et al., 2006; Nof et al., 2007; Rivolta et al., 2020; Schweizer et al., 2010; Ueda et al., 2004; Verkerk and Wilders, 2015). As mentioned, there has been important controversy regarding the role of HCN4 isoforms in generating human I_f_ and in pacemaking of sinoatrial node. There are reports demonstrating that the predominant HCN isoform at both the protein and RNA levels is HCN1 (Li et al., 2021, 2015). However, only a few data on the literature suggested the contribution of HCN1 variants to HR. One of them is a case report (Yu et al., 2021) showing two rare *HCN1* heterozygous variants identified in a young patient with profound sinus bradycardia. The second report identifies a polymorphic *HCN1* variant that was associated with HR variability and schizophrenia (Refisch et al., 2021). Conversely, dysfunctional HCN1 channels seem to play an important role in different forms of epilepsy and neuropathic pain (DiFrancesco et al., 2019; He et al., 2019). Our results add further support to the contention that in humans, HCN4 channels critically determine I_f_ density in sinoatrial cells, thus regulating HR.

The V240 residue is located in the N-terminus, and variants in this domain are less commonly described than in the pore and C-terminal regions of HCN4 channels. The HCND is formed by a stretch of 45 aminoacids directly preceding the S1 transmembrane segment that consists of three α-helices (HCNa, HCNb, and HCNc) (Figure 2) (Lee and MacKinnon, 2017). The V240 residue is actually located in the HCNc α-helix. The HCND could act as a sliding crank that converts the planar rotational movement of the CNBD into a rotational upward displacement of the VSD mechanically coupling thus, the CNBD and the VSD (Porro et al., 2019; Wang et al., 2020). HCND normally keeps the VSD in a position, which is unfavorable for channel opening (Porro et al., 2019). To test the possible molecular mechanism responsible for the gain-of-function effects of p.V240M we have mutated *in silico* a human HCN4 model (PDB:6GYN, Figure 12).

**Figure 12.**
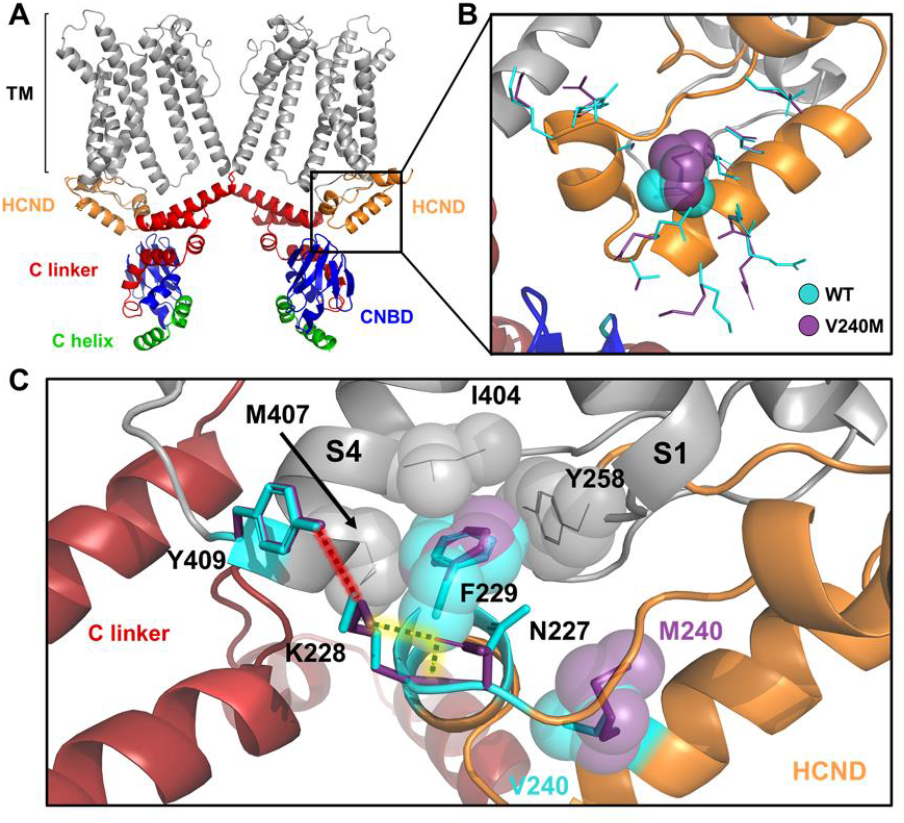
Molecular modeling of WT and p.V240M HCN4 channels. (**A**) Ribbon representation of the cryo-EM structure of human HCN4 (PDB:6GYN). Only two diagonal subunits are shown for clarity. The transmembrane segments (TM) are gray, the HCND is orange, the C-linker is red, the CNBD is blue, and the distal C helix is green. (**B**) Close view of superimposed WT and V240M models (colour coded as in **A**). Sticks representation shows amino acids side-chains from WT (cyan) and V240M (purple). V240 and M240 are shown as spheres. (**C**) Interaction of HCND with the VSD showing the hydrophobic interactions of F229 with I404 and M407 from TM S4 (VSD) and Y258 from the loop connecting HCND to TM S1, and polar contacts of K228 and Y409 in WT (red dashed line) or K228 and N227 in V240M (yellow dashed line). Spheres represent the van der Waals surface occupied by the side chains.

In HCN4 channels the HCND is anchored to the VSD by means of phenylalanine F229, whose aromatic side chain is inserted between transmembrane helices S1 and S4 in a hydrophobic pocket formed by I404 and M407 in S4, and Y258 in S1 (Figure 12B-C). Interestingly, the introduction of the methionine residue at this position remarkably reduced the energy in the most stable p.V240M conformation (–23.49 kcal/mol) compared to WT (–60.18 kcal/mol). Moreover, our molecular modeling demonstrates that the mutation introduces a slight torsion of the aromatic group of F229 and prevents the H-bond between K228 in HCND and Y409 in S4 (Figure 12C). These effects might weaken the interaction between HCND and VSD. Thus, these results suggest that p.V240M mutation limits the inhibitory action of the HCND on the VSD. The latter could explain the marked positive displacement (≈20 mV) of the activation curve of p.V240M channels. Indeed, our results are in accordance with those obtained by Porro et al (Porro et al., 2019) who showed that disrupting the interaction between the HCND and the S4 TM of the VSD produced a depolarizing shift in the voltage dependence of activation.

The cAMP-induced movement of the C-linker produced by the binding of cAMP to the CNBD is transmitted via the HCND to the VSD, lowering the energy barrier for channel opening (Porro et al., 2019; Wang et al., 2020). The mutants that disrupt the contact between the CNBD and the HCND eliminate the effect of cAMP on channel activation (Porro et al., 2019; Wang et al., 2020). Results of the molecular modeling demonstrated that the interaction between CNBD and HCND is not affected by the p.V240M mutation (Figure 12C). The latter would explain why p.V240M channels are sensitive to cAMP. Interestingly, our results demonstrated that p.V240M mutation markedly increases single HCN4 channel conductance, an effect that is beyond the positive shift of the activation curve. We propose that somewhat the mutation introduces changes in the pore region that controls the conductance of the channel, even when the channel selectivity is not affected. In summary, and considering that there are very few reports describing single-channel currents generated by HCN4 channels (Brandt et al., 2009; Michels et al., 2005), our results contribute to the understanding of the role of HCND in the distinctive biophysical properties of these channels.

The HCND is also necessary for the folding and trafficking of the channel to the plasma membrane (Porro et al., 2019; Wang et al., 2020). To the best of our knowledge, the only naturally occurring HCND mutation described so far is p.P257S which was identified in patients with early-onset atrial fibrillation (Macri et al., 2014). p.P257S channels present a trafficking defect being retained at the endoplasmic reticulum (Macri et al., 2014). Conversely, the p.V240M mutant is properly expressed in the membrane suggesting that p.V240M mutation does not affect the folding and trafficking of HCN4 subunits.

### Clinical implications

IST is a relatively common clinical entity producing highly distressing symptoms. In most cases, it is a sporadic non-familial-disease. Here we describe an autosomal-dominant form of IST with a striking complete penetrance. The clinical manifestations of the carriers are noteworthy, as they presented with systolic dysfunction. In fact, IST is barely associated with heart dysfunction and tachycardia-induced cardiomyopathy (Olshansky and Sullivan, 2019; Sheldon et al., 2015a) although some cases have been already reported (Romeo et al., 2011; Winum et al., 2009). Even when *HCN4* mutations have been related with noncompaction cardiomyopathy (Alonso-Fernández-Gatta et al., 2021), no signs of hypertrabeculation were found with CMR imaging or echocardiography in this family. The fact that ivabradine reversed systolic ventricular dysfunction suggests that the high HR and the duration of the arrhythmia (present from childhood but being documented even before birth in 2 patients) could have been the major contributors to cardiac dysfunction in our patients. Regarding treatment of IST, carriers were treated with ivabradine, since previous reports demonstrated that it reduced HR and improved quality of life in patients with IST (Cappato et al., 2012). Interestingly, the HR reduction in our study with ivabradine alone was ≈14% which is similar to that obtained previously in patients with IST of an unknown origin (Benezet-Mazuecos et al., 2013; Cappato et al., 2012). Responsiveness of our patients to ivabradine can be explained considering that sensitivity of p.V240M channels was identical to that of WT HCN4. These results strengthen the importance of genetic and functional studies for the design of personalized therapies in patients with inherited arrhythmogenic syndromes.

The degree of activation of I_f_ at the end of an AP determines the steepness of phase 4 depolarization; hence, the frequency of AP firing (Verkerk and Wilders, 2015). Thus, it seems reasonable to assume that the gain-of-function p.V240M mutation is responsible for an increase in sinoatrial node automaticity and the IST of the affected subjects in this family.

## Conclusions

Our results support the view that individuals carrying the p.V240M mutation present higher than normal I_HCN4_. This variant increases I_HCN4_ by an unprecedented mechanism that implies the alteration of the gating without modifying the channel sensitivity for cAMP or the membrane expression. Therefore, p.V240M is the first naturally occurring gain-of-function HCND variant which explains the faster than normal HR and IST of the carriers. The clinical, genetic and functional study of the variant led to the establishment of the personalized treatment with ivabradine for all carriers, contribute to shed light in the understanding of the pathophysiology of IST and help to the understanding the role of HCN4 channels in the pacemaker activity of human sinoatrial cells.

## Methods

### Study approval

All patients signed an informed consent, and the study was approved by the Research Ethics Committee of Granada (ref: CEIM/CEI/3-19 [12/4/19]. The purpose, methods and ethical aspects of this specific study were specifically assessed in an ordinary meeting of the Committee. Moreover, the study conforms to the principles outlined in the Declaration of Helsinki. Genetic results of the participants were reported following the guidelines proposed by the Working Group on Reporting Genetic Results in Research Studies (Bookman et al., 2006).

### Clinical study

The proband and relatives of a Spanish family with several members diagnosed with IST and left ventricular (LV) systolic dysfunction were evaluated at the **“***Hereditary cardiovascular disease unit***”** in the Cardiology Department of the Hospital Universitario Virgen de las Nieves. Trained personnel performed at least three electrocardiograms ECG at rest to the proband and to all the family members that gave their written informed consent. Family members either diagnosed or not with IST underwent a serial 24-hour ECG monitoring (at least two for patients diagnosed with IST). CMR was performed only to adult family members with IST. Three months after initiation of medical therapy with ivabradine (7.5 mg BID in adults, 5 mg BID in children) or ivabradine (5 mg BID) plus bisoprolol (5 mg BID), the 24-h Holter monitoring and echocardiogram were repeated for every patient, and the findings were compared with those obtained before treatment. The size of chamber, quantifications, and severity partition cut-offs of LV dysfunction were measured according to the current guidelines (Sheldon et al., 2015b). IST was defined as fast sinus rates (>100 bpm at rest or >90 bpm on average over 24 h) not due to underlying causes. Thus, prior to IST diagnosis, other causes such as hyperthyroidism, anemia, diabetes mellitus, orthostatic hypotension, infections, and drug abuse, were ruled out. A full pedigree was obtained collecting information such as cardiac events (sudden cardiac death, heart transplantation or device implantation), cases of IST, or systolic dysfunction of unknown etiology and any kind of cardiomyopathy across 4 generations. Echocardiographic, ECG, and imaging recordings were interpreted by experienced cardiologists, blinded to the genetic and clinical data of the participants. QT values were corrected using the Bazzet’s and Fredericia’s formulas in subjects with HR <100 and ≥100 bpm, respectively.

### Next-generation and Sanger sequencing

The genetic analysis of the proband was conducted by using a next-generation sequencing panel including 197 genes (Table Supplement File 1) following procedures previously described (Caballero et al., 2017; Nieto-Marín et al., 2022). Genomic DNA was extracted using an automated extraction and purification process by using the QIAsymphony SP® kit (Qiagen, Hilde, Germany) according to the manufacturer’s instructions. Library preparation was carried out using SureSelect Reagent kit (Agilent, Santa Clara, CA, USA) for Illumina’s paired-end multiplexed sequencing method, following the manufacturer’s instructions. The enrichment of the regions of interest was performed by means of a SureSelect (Agilent) probe kit that selectively captures the coding zones and the flanking intronic areas of the selected genes. After the generation of clusters, the DNA libraries were sequenced on the Illumina HiSeq 1500 platform. The analysis of the sequencing data was performed using a proprietary bioinformatics pipeline to obtain a report of variants noted along with their coverage and corresponding quality parameters. We excluded variants located in introns (except those in suspected splicing sites) or intergenic regions. We also removed synonymous variants and non-synonymous variants with occurrences >1 in our local database. The list of variants identified were evaluated against database information on previously described variants [Human Gene Mutation Database (http://www.hgmd.cf.ac.uk/), Single Nucleotide Polymorphism (SNP) database (http://www.ncbi.nlm.nih.gov/SNP/), NHLBI GO Exome Sequencing Project (http://evs.gs.washington.edu/EVS), ClinVar (https://www.ncbi.nlm.nih.gov/clinvar/), or Genome Aggregation database (http://gnomad.broadinstitute.org). Pathogenicity of the identified variants was established according to the current recommendations of the American College of Medical Genetics and Genomics and the Association for Molecular Pathology (Richards et al., 2015). Variant pathogenicity was graded according to its presence in a previously associated or candidate gene and the *in silico* predicted impact on the protein using widely used software (PROVEAN, SIFT, Polyphen2, Mutation Taster, and Combined Annotation Dependent Depletion Score), the degree of conservation of the affected residue measured by multiple ortholog alignment using Alamut software (http://www.interactive-biosoftware.com).

A phenotype-genotype segregation study was conducted through cascade screening among available relatives with Sanger genetic study (Caballero et al., 2017; Nieto-Marín et al., 2022). PCR products were purified using illustra ExoProStar 1-Step (GE Healthcare Life Sciences, Chicago, IL, USA) and the analysis was performed by direct sequencing (Health in Code, A Coruña, Spain). The results were compared with the reference sequence from hg19 by means of Chromas Lite Software (http://technelysium.com.au).

### Cell culture and transfection

The study has been conducted in CHO and HEK-293 purchased from American Type Culture Collection (ATCC, Manassas, VA, USA). They had been authenticated by the supplier as appropriate. Mycoplasma tests were conducted routinely for both cell lines and showed no mycoplasma contamination.

CHO cells were grown in Ham-F12 medium supplemented with 10% fetal bovine serum, 100 U/ml penicillin, and 100 μg/ml streptomycin (Caballero et al., 2010; Caballero et al., 2017; Pérez-Hernández et al., 2018; Alonso-Fernández-Gatta et al., 2021; Crespo-García et al., 2023). HEK-293 cells were cultured in Dulbecco’s modified Eagle’s (DMEM) medium supplemented with 10% fetal bovine serum, 100 U/ml penicillin, and 100 μg/ml streptomycin as previously described (Caballero et al., 2010; Crespo-García et al., 2023). The cultures were passed every 2–5 days using a brief trypsin treatment.

The p.V240M HCN4 mutation (NP_005468.1) was introduced by using the QuikChange Site-Directed Mutagenesis kit (Agilent, Santa Clara, CA, USA) and confirmed by direct DNA sequencing (Secugen S.L., Madrid, Spain). The sequence of the oligonucleotides used to introduce the p.V240M substitution was the following: sense 5’ GCAGCCAGAAAGCCaTGGAGCGCGAACAG 3’; antisense 5’ CTGTTCGCGCTCCAtGGCTTTCTGGCTGC 3’. The sequence of the oligonucleotide used for sequencing was: 5’ ACGGACACCTGCATGACTC 3’.

For macroscopic current recordings, subconfluent cultures were transiently transfected with cDNA encoding WT or mutated HCN4 channels (1.6 µg) together with cDNA encoding the CD8 antigen (0.5 µg) by using X-tremeGENE™ HP (Roche Diagnostics, Rotkreuz, Switzerland) in CHO cells or Lipofectamine 2000 (Thermofisher Scientific, Waltham, MA, USA) in HEK-293 cells, with manufacturer instructions followed (Caballero et al., 2010; Caballero et al., 2017; Pérez-Hernández et al., 2018; Alonso-Fernández-Gatta et al., 2021; Crespo-García et al., 2023). In some experiments, WT and p.V240M HCN4 were cotransfected (WT+p.V240M) at a 0.5:0.5 ratio (0.8 µg each). For single channel recordings, CHO cells were transfected with 0.5 instead of 1.6 µg of the cDNA encoding mutated or WT HCN4.

In all cases, at the point of 48 h after transfection, cells were incubated with polystyrene microbeads precoated with anti-CD8 antibody (Dynabeads M450, Thermofisher Scientific). Most of the cells that were beaded also exhibited channel expression. The day of recordings, cells were removed from the dish by using a cell scraper or by trypsinization, respectively, and the cell suspension was stored at room temperature and used within 12 h for electrophysiological experiments.

To minimize the influence of the expression variability, each construct was tested in a large number of cells obtained from at least 3 different cell batches. Moreover, to avoid putative interferences of culture conditions (passage number, cell density, etc), currents generated by cells expressing WT, p.V240M and WT+p.V240M channels were always recorded in parallel.

### Recording techniques

#### Macroscopic current recordings

A small aliquot of cell suspension was placed in a 0.5 ml chamber mounted on the stage of an inverted microscope (Nikon TMS; Nikon Co., Tokio, Japan). After settling to the bottom of the chamber, cells were perfused at ≈1 mL/min with external solution (see the composition below). Macroscopic currents were recorded at room temperature (21-23ºC) by means of the whole-cell patch-clamp technique using Axopatch-200B patch clamp amplifiers and pCLAMP software (Molecular Devices, San José, CA, USA) (Caballero et al., 2010; Caballero et al., 2017; Pérez-Hernández et al., 2018; Alonso-Fernández-Gatta et al., 2021; Nieto-Marín et al., 2022; Crespo-García et al., 2023) Recording pipettes were pulled from 1.0 mm o.d. borosilicate capillary tubes (GD1, Narishige Co., Ltd, Tokio, Japan) using a programmable patch micropipette puller (Model P-2000 Brown-Flaming, Sutter Instruments Co., Novato, CA, USA) and were heat-polished with a microforge (Model MF-83, Narishige). Micropipette resistance ranged 3-5 MΩ when filled with the internal solution and immersed in the external solution. In all the experiments, series resistance was compensated manually by using the series resistance compensation unit of the Axopatch amplifier, and ≥80% compensation was achieved. The remaining access resistance after compensation and cell capacitance were 1.7±0.4 MΩ and 11.9±0.7 pF (n=65), respectively. Therefore, under our experimental conditions no significant voltage errors (<5 mV) due to series resistance were expected with the micropipettes used. Currents were filtered at half the sampling frequency and stored on the hard disk of a computer for subsequent analysis. CHO cells were perfused with an external solution containing (mM): NaCl 110, KCl 30, HEPES 5, MgCl_2_ 0.5, CaCl_2_ 1.8, and glucose 10; pH= 7.4 with NaOH. Recording pipettes were filled with an internal solution containing (mM): K-aspartate 80, KCl 42, KH_2_PO_4_ 10, MgATP 5, phosphocreatine 3, HEPES 5 and EGTA 5; pH 7.2 with KOH. The protocol to obtain current-voltage relationships consisted of 2-s steps that were imposed in 10 mV increments from a holding potential of -40 mV to potentials ranging –160 and 0 mV. Current amplitude was measured at the end of the pulse and normalized in each experiment to membrane capacitance to obtain current densities. To determine putative effects on the ion selectivity of HCN4 channels, the reversal potential (E_rev_) was measured at extracellular and intracellular K^+^ concentrations of 30 and 142 mM, respectively. To this end, 3-s pulses to –120 mV followed by 1-s pulse to membrane potentials ranging –50 to +20 mV were applied. The E_rev_ was calculated in each experiment from the intersection of the linear regression to the data with the abscissas axis. To analyze potential effects on the time course of current activation, a monoexponential function was fitted to the activation phase of current traces elicited by pulses to –160 mV yielding τ_act_ that defines the process. To measure HCN4 deactivation kinetics, two 3-s pulses from –40 to –120 mV were applied at increasing coupling intervals (1-6 s) (Mistrík et al., 2006; Schweizer et al., 2010). This protocol allowed us to calculate, for each coupling interval, the ratio between ΔI_INS_ and I_SS_ elicited at the beginning and at the end, respectively, of the second pulse to –120 mV. The calculated ratio was plotted as a function of the coupling interval and a monoexponential function was fitted to the data to obtain the τ_deact_.

The protocol to analyze the voltage dependence of HCN4 channel activation consisted of 2-s pulses from –40 mV to potentials ranging from –160 to 0 mV followed by 1-s pulses to –140 mV to record the tail currents. The tail current amplitude was normalized to the maximum value and plotted as a function of the membrane potential of the preceding pulse to construct the activation curves. A Boltzmann function was fitted to the data to calculate the midpoint (V_h_) and the slope (*k*) of the curves. In a subset of experiments, the consequences of the p.V240M variant on the shift of the voltage-dependent activation produced by of 3’,5’-cyclic Adenosine monophosphate (cAMP) were analyzed in HEK-293 cells. It has been recently described that in CHO but not in HEK-293 cells, HCN4 channel activation is constitutively shifted to more depolarized membrane potentials and is no longer affected by cAMP (Peters et al., 2020). In these experiments, HEK-293 cells cultured and transfected as described above, were perfused with the same external solution than described for CHO cells (see above), while the internal solution contained (mM): KCl 130, NaCl 10, MgATP 2, HEPES 5, MgCl_2_ 0.5 and EGTA 1; pH 7.2 with KOH. cAMP (Merck, Rahway, NJ, USA) at a saturating concentration (10 μM) was added to the pipette solution.

The effects of different concentrations of ivabradine (0.1-3 µM) was determined in CHO cells transfected with WT or p.V240M HCN4 channels by applying trains of activating/deactivating steps to -140 and +5 mV from a holding potential of -35 mV (Bucchi et al., 2013). The I_HCN4_ inhibition at –140 mV was used as an index of block (*f;* expressed as percentage of inhibition) to construct the respective concentration-response curves and a Hill equation (*f* = 1/{1+(IC_50_/[D])^nH^}) was fitted to the data to obtain the concentration needed to produce the 50% of the maximum inhibition (IC_50_). In the fit, n_H_ was fixed to unity and bottom and top values were constrained to 0 and 100%, respectively.

#### Single channel recordings

Single channel currents were recorded in CHO cells at room temperature (21-23ºC) using the cell-attached patch-clamp configuration (Michels et al., 2005; Brandt et al., 2009; Caballero et al., 2010; Crespo-García et al., 2023). Using this configuration, the intracellular environment is completely preserved and channel activity can be measured in an intact cell. Cells were suspended in bath solution containing (mM): KCl 130, NaCl 10, EGTA 5 and HEPES 10; pH 7.4 with KOH. This high-K^+^ solution was used to achieve a resting membrane potential of zero. Patch pipettes were pulled from 1.5 mm o.d. borosilicate capillary tubes (Harvard Apparatus Ltd, Holliston, MA, USA), coated at the tip with Sylgard (Dow Corning, Midland, MI, USA), and fire-polished with a microforge (Mod. MF-830. Narishige). When filled with pipette solution containing (in mM): KCl 70, NaCl 70, MgCl_2_ 1, BaCl_2_ 2 and HEPES 5; pH 7.4 with KOH, tip resistances were between 5 and 10 MΩ. The micropipettes were gently lowered onto the cells to get a gigaohm seal after applying suction. After seal formation, the cells were lifted from the bottom of the perfusion bath and current recordings were started. Single channel currents were recorded by applying repetitive 10-s pulses from a holding potential of –35 mV to –90 mV or to potentials between –120 and +40 mV in 10 mV steps. Current data were sampled at 10 kHz and filtered at 1 kHz. The apparent number of active channels in a patch was determined by visual inspection of the current traces and patches with more than one channel were discarded. The experimental conditions were optimized to reduce the number of active channels on each recording: e.g., the amount of cDNA used for cell transfection was reduced by ≈70% and the tip resistance of the pipettes was increased approximately threefold compared to whole-cell recordings. Under these conditions, only 10% of cells with active channels had more than one channel. All data analysis was performed by using pCLAMP software and opening events were captured using the event detection tool of Clampfit 10 (Caballero et al., 2010; Crespo-García et al., 2023). At the end of each experiment, the HCN channel inhibitor ivabradine (5 µM) (Bucchi et al., 2006) was added to the external solution to specifically abolish single channel currents generated by HCN4 channels.

Amplitude of unitary currents was determined for each experiment by direct measurements of fully resolved openings, which allowed the calculation of average values. The opening probability (*P*_o_) was obtained for each experiment by dividing the time that the channel remains in the open state by the total recording time and the opening frequency (*f*_o_) was calculated for each experiment as the inverse of the closed time between events. The mean open time was calculated for each experiment by dividing the time the channel remains in the open state by the events. To measure opening and closing kinetics, dwell-time histograms were constructed by plotting pooled dwell-time data as a function of the number of events per bin that were fitted by monoexponential functions to calculate τ_OPEN_ and τ_CLOSED_. Current–voltage relationships were constructed by plotting the single channel current amplitude as a function of the membrane potential and conductance (γ) was calculated from the slope of the fit of a linear function to the data recorded at potentials between –120 and –60 mV.

#### Biotinylation assay

A biotinylation assay was conducted in CHO cells using procedures previously described (Crespo-García et al., 2023). At the point of 48 h after transfection of HCN4 WT or p.V240M, CHO cells were washed twice with ice cold phosphate-buffer saline (PBS) and biotinylated for 15 min at 4°C using PBS containing 0.5 mg/mL of EZ Link Sulfo-NHS-SS-Biotin (ThermoFisher). Plates were washed twice with PBS-200 mM glycine (to quench unlinked biotin) and again with PBS. Cells were collected in RIPA buffer containing 50 mM Tris·HCl (pH=7.5), 150 mM NaCl, 1% Nonidet P-40, 0.1% SDS, 0.5% sodium deoxycholate, and 1 mM phenylmethylsulfonyl fluoride (PMSF) and protease inhibitor cocktail (Merck) for protein extraction. Subsequently, the extract (1 mg) was incubated with Streptavidin Sepharose (50 µl, GE Healthcare) overnight at 4°C. To separate biotinylated fraction, samples were centrifugated at 3000 rpm for 2 min at 4°C. The fraction of biotinylated proteins was washed by several centrifugations. Thereafter, HCN4 and ezrin proteins were detected by Western blot following previously described procedures (Pérez-Hernández et al., 2018; Nieto-Marín et al., 2022; Crespo-García et al., 2023). Nuclei and cell debris were removed by centrifugation at 14.000 rpm for 20 min at 4°C. The total protein amount of the extracts was calculated with the bicinchoninic acid method (Pierce™ BCA Protein Assay Kit, ThermoFisher). Samples were run on 4-15% Mini-PROTEAN TGX™ stain-free gels (Bio-Rad, Hércules, CA, USA USA) and, afterwards, protein was transferred to nitrocellulose membranes. Nonspecific binding sites were blocked with 5% non-fat dried milk in PBS with Tween-20 (0.05%) for 1 hour at room temperature. Membranes were then incubated with rabbit polyclonal anti-HCN4 (1:1000; 55224-1-AP, Proteintech, Rosemont, IL, USA) and mouse monoclonal anti-ezrin (1:400; ab4069, Abcam, Cambridge, UK) primary antibodies overnight at 4°C. All the antibodies had been validated by the manufacturers. After the incubation with the primary antibodies, samples were incubated for 1 hour with peroxidase-conjugated goat anti-rabbit (111-035-144) or goat anti-mouse (115-035-003) secondary antibody (1:10000; Jackson Immunoresearch, West Grove, PA, USA). Membranes were washed three times with PBS-Tween after adding primary and secondary antibodies. Protein expression was detected by chemiluminescence (SuperSignal™ West Femto Maximum Sensitivity Substrate, ThermoFisher) and visualized using the Chemidoc MP System and Image Lab 5.2.1. software (Bio-Rad). Expression of the proteins in the biotinylated (membrane) fraction was normalized to the input expression.

#### Mathematical modeling of human sinoatrial node action potential

We employed the Fabbri-Fantini-Wilders-Severi model of human sinus node cells (Fabbri et al., 2017). The model is based on experimental recordings and numerical reconstructions of the I_f_ measured in human sinoatrial node cells (Verkerk et al., 2007a, 2007b) and fairly reproduces the changes in heart rate induced by mutations or drugs affecting the I_f_ (Fabbri et al., 2017). We downloaded the model files available at the CellML model repository (http://models.cellml.org/cellml), as part of the International Union of Physiological Sciences Physiome Project (Yu et al., 2011). To perform the simulations we used OpenCell software package that allows reading, modifying, solving, and plotting CellML models (http://cellml-opencell.hg.sourceforge.net:8000/hgroot/cellml-opencell/cellml-opencell). Initially, the model was run (for 100 s) keeping all parameters at their default values and under these conditions, the model was stable and reproduced the results obtained in the original description of the model (Fabbri et al., 2017). The results obtained with these “basal” conditions were considered to correspond to WT HCN4. Thereafter, the experimentally measured changes produced by p.V240M HCN4 or WT + p.V240M HCN4 on the conductance, kinetics and voltage dependence of activation of the macroscopic I_f_ were incorporated into the model equations corresponding to the I_f_. The modified versions of the model were also run for 100 s. In all cases, we plotted action potential and I_f_ traces yielded at the end of the simulation runs and several parameters were measured including cycle length, beating frequency, diastolic depolarization rate, mean diastolic potential, action potential amplitude, overshoot, and action potential duration measured at 20, 50, and 90% of repolarization.

### Molecular modeling

To estimate the conformational changes produced by the p.V240M HCN4 mutation a molecular modeling using the crystallized structure of human HCN4 (PDB: 6GYN) was performed. Model refinement, in silico mutagenesis, and energy calculation were done with FoldX Suite (version 4.0, https://www.foldxsuite.crg.eu). For model refinement and minimizing the energy of the structure, the RepairPDB protocol was used. The Val-to-Met substitution at position 240 of each monomer of the optimized PDB was performed and the protocol to calculate free energies was run ten times. Default parameters of the software were used except temperature, pH, ionic strength and VdW design that were set at 298K, 7, 5 and 0, respectively. When the model running was completed, the software provided different conformations with different energies and the conformation of the mutated form with the lowest energy was chosen. The optimized conformations of WT and p.V240M HCN4 were compared on PyMOL (Molecular Graphics System, Version 2.0 Schrödinger, LLC; https://www.pymol.org/2/) and putative changes in the aminoacid positions and interactions (Hydrophobic, electrostatic and H-Bonds) with surrounding residues induced by the presence of the mutation were identified (Caballero et al, 2010).

### Statistical Analyses

Clinical and experimental results are expressed as mean[SD] and mean±SEM, respectively. Small-sized samples were first analyzed to determine the distribution of the variables (Normality) using the Shapiro–Wilk and Kilmogorov-Smirnov tests. When these tests demonstrated a normal distribution, parametric tests were used. Paired or unpaired *t* test, multiple *t* test, one-way ANOVA followed by Tukey’s test, or two-way ANOVA were used to assess statistical significance where appropriate. When a statistically significant difference was determined by these tests, a Pearson correlation was performed to confirm that the observed differences were not due to random sampling. To make comparisons between fits of pooled or of concentration-dependent data, an F-test was used. Variance was comparable between groups throughout the manuscript. A value of *P*<0.05 was considered significant. Statistical tests were conducted using GraphPad Prism 8 (https://www.graphpad.com/scientific-software/prism/) or Microsoft Excel. For the different groups of experiments sample size was chosen empirically according to previous experience in the calculation of experimental variability. No statistical method was used to predetermine sample size. The cellular experiments were not blinded due to the nature of the experimental design and platforms but all the data were analysed in an identical manner for all conditions to eliminate possible operator bias.

## Data Availability

All data generated or analyzed during this study are included in the manuscript and supporting files. CMR and acute and Holter ECGs recordings will be not available since they are incorporated into the confidential medical record of each participant.

